# Mental health, blood pressure and the development of hypertension

**DOI:** 10.1101/2022.11.04.22281936

**Authors:** H. Lina Schaare, Maria Blöchl, Deniz Kumral, Marie Uhlig, Lorenz Lemcke, Sofie L. Valk, Arno Villringer

**Affiliations:** Department of Neurology, Max Planck Institute for Human Cognitive and Brain Sciences, Leipzig, Germany; Institute for Psychology, Leipzig University, Leipzig, Germany; MindBrainBody Institute, Berlin School of Mind and Brain, Berlin, Germany; Nuclear Magnetic Resonance Unit, Max Planck Institute for Human Cognitive and Brain Sciences, Leipzig, Germany; Clinic of Cognitive Neurology, Leipzig University, Leipzig, Germany; Charité University Medicine Berlin, Berlin, Germany; Otto-Hahn-Group Cognitive Neurogenetics, Max Planck Institute for Human Cognitive and Brain Sciences, Leipzig, Germany; Institute of Neuroscience and Medicine (INM-7: Brain and Behaviour), Research Centre Jülich, Germany; Institute of Psychology, Neuropsychology, University of Freiburg, Freiburg, Germany; Institute of Psychology, Clinical Psychology and Psychotherapy Unit, University of Freiburg, Freiburg, Germany; Institute of Systems Neuroscience, Medical Faculty, Heinrich Heine University Düsseldorf, Düsseldorf, Germany

## Abstract

Hypertension (HTN) has been associated with a greater risk of affective disorders. Paradoxically, several studies have shown the opposite effect in which high blood pressure relates to less depressive symptoms and greater well-being. Here we dissolve this paradox and clarify the relationship between mental health, blood pressure and the development of HTN using the UK Biobank. In adjusted multiple linear regression models, we found that the presence of a HTN diagnosis was associated with impaired mental health (i.e. more depressive symptoms (N = 303,771; β = 0.043; 95% CI [0.039, 0.047]; p<0.001) and lower well-being scores (N = 129,876; β = -0.057; 95% CI [-0.064, - 0.050]; p<0.001)) at baseline, whereas higher systolic blood pressure (SBP) was associated with fewer depressive symptoms (N = 303,771; β = -0.063; 95% CI [-0.067, -0.060]; p<0.001) and higher well-being scores (N = 129,876; β = 0.057; 95% CI [0.051, 0.063]; p<0.001). These effects persisted until follow-up (∼10 years later). To explore a potential link between the mental health-blood pressure association and the development of HTN, we compared participants who were normotensive at baseline and developed HTN until follow-up with those who stayed normotensive. Notably, the adjusted model showed impaired mental health already at baseline in HTN developers (i.e., before HTN diagnosis; depressive symptoms: β = 0.060; 95% CI [0.045, 0.076]; p<0.001; well-being: β = -0.043; 95% CI [-0.068, -0.017]; p<0.001), indicating that people who develop HTN might require higher blood pressure levels for the same mental health outcomes as normotensives. In addition, the negative association between SBP and depressive symptoms at baseline was moderated by HTN development (β = -0.014; 95% CI [-0.026, -0.003]; p=0.015), suggesting that the negative relationship between mental health and blood pressure was accentuated in people developing HTN several years before receiving their HTN diagnosis. We further observed that higher SBP was associated with lower emotion-related brain activity from functional magnetic resonance imaging (fMRI; β = -0.032 95% CI [-0.045, -0.019]; p<0.001). This effect was also moderated by HTN diagnosis, suggesting an impact of SBP and HTN on the central nervous processing of emotions. Possible mechanisms are discussed, including regulatory baroreceptor circuits linking arterial blood pressure to neural processing of emotions. Overall, our results show an interrelation between mental health and blood pressure that may be involved in the development of HTN. In people who develop HTN, this relationship seems to be altered, such that higher blood pressure is required to sustain mental health, potentially offering a novel perspective for developing preventive and therapeutic measures.

## Introduction

Both hypertension (HTN) and affective disorders, such as depression, frequently co-occur and have been identified as single ^1–4^ as well as combined ^5^ risk factors for cardiovascular disease (CVD). An increased risk of HTN has been described in patients with affective disorders ^6–10^. The burden of vascular risk factors, including HTN, has further been suggested to drive depressive symptoms in ageing through microvascular brain damage ^11^.

In contrast to these findings, some studies showed that higher blood pressure related to better mood, higher well-being and lower distress in healthy ^12–16^ and clinical populations ^17–19^. Baroreceptor mechanisms have been suggested to explain these effects, as their intrinsic and experimentally-induced signalling has been shown to phasicly adjust pain sensitivity thresholds, alter sensory and emotional processing, decrease cortical excitability and inhibit central nervous system activity ^20–26^. These observations have been proposed as a critical neuro-behavioural component in the development of essential HTN. Momentary relief from an adverse state might positively reinforce blood pressure-elevating behaviours and thus, via baroreceptor-mediated neural circuits, insidiously increase blood pressure over time, resulting in ‘learned hypertension’ ^20,25,27–29^. However, it remains unclear if blood pressure elevations and HTN development relate to mental health and if such an association reflects in brain function.

The first goal of the present study was to systematically describe the relationship of blood pressure with depressive symptoms and well-being, while accounting for potential confounding effects of medication intake and chronic illness, such as CVD and clinical depression. Due to small effect sizes reported in previous research related to our study ^16–18^, we capitalized on the unique study design offered by the UK Biobank. The UK Biobank combines a deeply phenotyped longitudinal cohort with high statistical power of more than 500,000 participants ^30^ which enables the detection of robust small effects. In addition, it includes two follow-up timepoints for longitudinal analyses in two sub-samples of the baseline cohort: an online mental health follow-up at around 5 years and a follow-up at around 10 years. We hypothesized for cross-sectional and longitudinal analyses that increased blood pressure relates to fewer depressive symptoms and greater well-being (preregistration: https://osf.io/638jg/).

The second goal of this study was to explore the relevance of blood pressure-mental health associations in relation to HTN development. If a systematic relationship between mental health and blood pressure exists, this could positively reinforce long-term blood pressure elevations via baroreceptor mechanisms. Hence, we explored if the relationship between mental health and blood pressure differed between participants who developed hypertension compared to those who stayed normotensive.

The third goal of this study was to explore the prospective effects of blood pressure-mental health associations in emotion-related brain function using UK Biobank’s imaging assessment (N > 20,000). Due to the previously established baroreceptor effects on central nervous processing, blood pressure elevations and the development of HTN might relate to more general affective processes in brain function. We therefore investigated the effect of blood pressure variations on emotional task-based functional MRI activation ^31,32^.

## Methods

This study was conducted using data from the UK Biobank (http://www.ukbiobank.ac.uk/) to investigate cross-sectional and longitudinal associations between blood pressure, hypertension and mental health in behaviour and brain. We were granted access to UK Biobank’s data resource in May 2018 after an initial application, but embargoed data access until we completed the preregistrations of this and related studies (date of initial data release 12 Feb 2019, preregistration of this study: https://osf.io/638jg/). All analyses and data visualisations were performed with R (4.0.2, R Core Team, 2015, Vienna, Austria; R-project.org/). To allow for replication studies and reproducibility of our results, the analysis code can be found in the Open Science Framework repository of this study (https://osf.io/638jg).

### UK Biobank study design

The UK Biobank is a publicly available, on-going longitudinal study which aims to comprehensively assess health-related indices of more than 500,000 UK citizens ^30^. UK Biobank included participants from the UK between the ages of 40 and 69 years at recruitment in 2006 to 2010. The size of the cohort was determined based on statistical power calculations for nested case-control studies to achieve large incidences and reliable odds ratios of common health-related conditions during the first years of the 20-years follow-up period ^30^.

The initial assessment of the whole cohort was conducted between 2006-2010 in 22 assessment centres throughout the UK. The complete assessment protocol was repeated at two other instances, specifically at first repeat assessment visit (2012-2013), and at imaging follow-up which included ongoing brain magnetic resonance imaging (MRI) assessments of 100,000 UK Biobank participants that has started in 2014 ^33,34^. In addition, a sub-sample of the baseline cohort participated in an online mental health follow-up assessment in 2016. For this study, we used data from the initial assessment visit (i.e., the baseline visit), the online mental health follow-up at around 5 years from baseline, as well as the follow-up imaging visit (i.e., the follow-up visit) at around 10 years from baseline (Figure 1).

**Figure 1.**
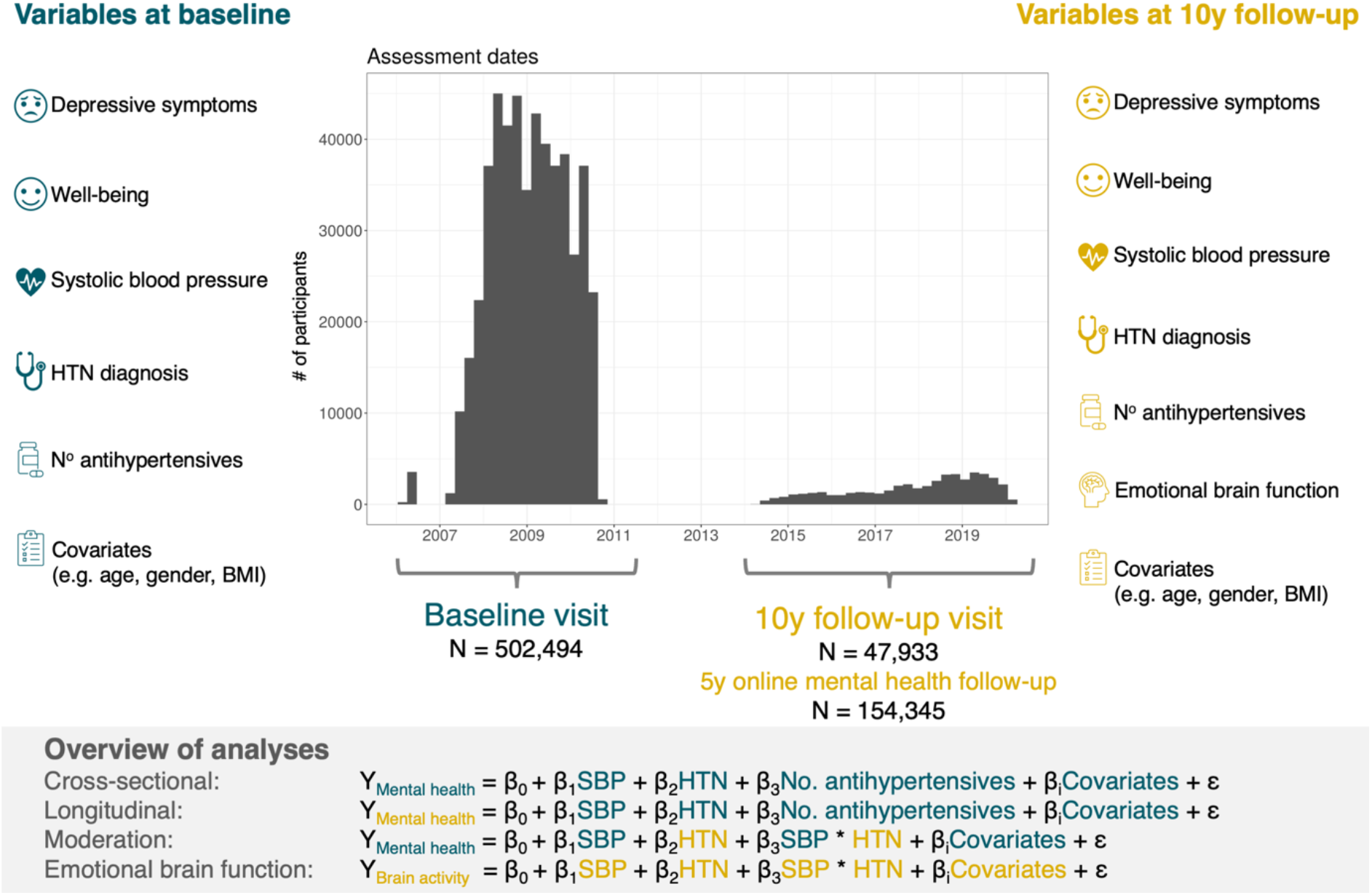
Overview of study design, outcome and predictor variables and analyses.

### Variables

A set of questions and measures designed for the assessment of current and lifetime mental health and psychosocial factors were administered at several instances during UK Biobank data acquisition ^35–37^. The following variables were included in this study (details on the UK Biobank data fields that were used for each variable are reported in the Supplementary Materials):

#### Outcomes

##### Current depressive symptoms

During each assessment centre visit, frequency of current depressive symptoms in the last two weeks (i.e., depressed mood, unenthusiasm, tenseness, tiredness) was assessed using a 4-point Likert scale ranging from 0 (“not at all”) to 3 (“nearly every day”). The item scores were summarized as mean scores for the purpose of this study. Questions regarding current depressive symptoms were assessed via a touchscreen during baseline and follow-up assessment centre visits ^35–37^.

We also preregistered analyses using the Patient Health Questionnaire 9-question version (PHQ-9) ^38^ from the online mental health follow-up assessment which a sub-sample of the whole UK Biobank cohort received ^35^. In the online PHQ-9 questionnaire, severity of current depressive symptoms in the last two weeks was assessed. The analyses and results of this questionnaire were almost identical to the ones obtained from the main depressive symptoms outcome measure described above. Methods and results related to PHQ-9 are further reported in the Supplementary Materials.

##### Well-being

Seven questions addressing different aspects of participants’ well-being were included at the assessment centre visits. The questions asked how happy/satisfied participants were regarding their happiness, and health, work, family, friendship, and financial situation, respectively. Participants were asked to respond to the questions on a 6-point Likert scale ranging from 1 (“extremely happy”) to 6 (“extremely unhappy”). For the purpose of this study, all item scores were recoded and summarized as mean scores with higher scores representing greater well-being.

#### Predictors

##### Systolic blood pressure (SBP)

Systolic and diastolic blood pressure readings were taken from all UK Biobank participants. At each assessment centre visit, two readings were recorded with an automated Omron 705 IT electronic blood pressure monitor (OMRON Healthcare Europe B.V. Kruisweg 577 2132 NA Hoofddorp). The two blood pressure readings were obtained during seated resting periods interleaved with a verbal interview on demographic and health-related data by UK Biobank staff. The procedure was conducted in the following sequence: 1) Interview Part 1 (demographic data), 2) First measurement of blood pressure, 3) Interview Part 2 (diseases and medications), 4) Measurement of Pulse Wave Velocity, 5) Second measurement of blood pressure. A timer ensured the second blood pressure reading could not be taken until at least 1 minute had elapsed. We used the mean of the two readings to derive systolic (SBP) values per participant at baseline and follow-up visit.

##### Hypertension diagnosis (HTN)

Complementary to blood pressure readings, we used a categorical measure of HTN diagnosis. Participants indicated on a touchscreen whether a doctor has ever told them that they have had high blood pressure. These self-reports were given at each assessment visit and used in this study as an assessment if a diagnosis of HTN was present (yes/no) at baseline and follow-up.

##### Number of antihypertensive medications

In a nurse interview at each assessment visit, participants reported all medications they were currently taking. A physician from our team (LL) examined these medication lists and identified all antihypertensive drug classes (Supplementary Materials). We subsequently coded these antihypertensives and calculated a sum score for each participant, indicating the number of antihypertensive drugs taken. A higher number of antihypertensive medications suggests that different drug classes are required to counteract the effects of blood pressure dysregulation and thus serves as an indicator of HTN severity.

#### Additional variables used in exploratory analyses

##### Definition of hypertension

In the moderation analysis (see below), we aimed to explore the relevance of blood pressure-mental health associations in relation to HTN development. For this purpose, our approach was to define HTN as conservatively as possible to avoid 1) missing hypertensive cases and 2) confounding influences of antihypertensive intake. Hence, we defined hypertension for this analysis as either having a HTN diagnosis (as described above) or using any antihypertensive medication. For the moderation analysis, this definition was used for exclusion of participants with hypertension at baseline to select a non-hypertensive sample at baseline. At follow-up, this definition was used to define which participants became hypertensive between baseline and follow-up (i.e. having a HTN diagnosis at follow-up or taking antihypertensives at follow-up) and who stayed normotensive.

##### Imaging-derived phenotypes (IDPs)

For analyses relating to brain function, we utilised the imaging-derived phenotypes (IDPs) available in UK Biobank which have been generated by processing the raw neuroimaging data. Magnetic resonance imaging (MRI) was performed at 3 Tesla and details of the imaging protocols and data processing procedures have been described in detail elsewhere ^33,34^. In brief, the functional MRI task that was implemented in UK Biobank is the Hariri “emotion” task ^31,32^. Participants were presented with blocks of trials and asked to decide either which of two faces presented on the bottom of the screen match the face at the top of the screen, or which of two shapes presented at the bottom of the screen match the shape at the top of the screen. The faces had either an angry or fearful expression. Trials were presented in blocks of 6 trials of the same condition (faces or shapes), with the stimulus presented for 2000 ms and a 1000 ms inter-trial interval. Each block was preceded by a 2000 ms task cue (“shape” or “face”), so that each block was 21 seconds including the cue. Each of the two runs included 3 face blocks and 3 shape blocks, with 8 seconds of fixation at the end of each run. IDPs of the Hariri task reflect the strength of response to the stimuli within a given brain region using the blood oxygenation level dependent (BOLD) contrast. BOLD fMRI relies on regional differences in haemoglobin oxygenation (more precisely local concentration changes of deoxy-haemoglobin) to measure regional brain activity ^39–41^. Here, we focused on the IDPs of significant clusters in amygdala and whole-brain activation for the faces > shapes contrast (median z-statistics of BOLD activation), which reflects the emotional brain response.

#### Covariates

To adjust for confounding factors, we included the following variables as covariates in the analyses: age, gender, body mass index (BMI), resting heart rate, diabetes diagnosed by doctor (yes/no), lifetime depression diagnosed by doctor (yes/no), angina diagnosed by doctor (yes/no), myocardial infarction diagnosed by doctor (yes/no). Note that we refer to the variable *gender* and not sex, as the UK Biobank defines this variable as a mixture of the sex the NHS had recorded for the participant and self-reported sex, although we acknowledge that this does not capture the full spectrum of gender.

### Statistical methods

We performed cross-sectional and longitudinal multiple linear regression models. For cross-sectional comparisons, we used data of the initial assessment visit. Outcomes were current depressive symptoms and well-being, respectively. SBP, HTN and the number of antihypertensives were entered simultaneously as predictors. For longitudinal comparisons, we predicted outcome measures (i.e. depressive symptoms and well-being) assessed at the follow-up visit from predictors assessed at the initial assessment visit (i.e. SBP, HTN and the number of antihypertensives). The analyses were performed for the total sample, as well as for a subset of participants with HTN diagnosis. All statistical models included the same covariates from initial assessment visit (specified above, i.e., age, gender, BMI, resting heart rate, history of diabetes, angina, myocardial infarction, and lifetime depression). Missing data were listwise excluded and outcomes, predictors and covariates were z-scored. In all models, we have also assessed any potential influence of multicollinearity by evaluating the Variance Inflation Factor (VIF). The VIFs never exceeded a value of 2, which indicates that multicollinearity is low and inferences from our models are likely not biased due to correlations among the variables included.

#### Sensitivity analyses

We investigated the sensitivity of effects by inclusion and exclusion of participants with diseases which often affect blood pressure and/or mental health. As such, we considered diagnoses of HTN, non-bipolar depression, and severe neurological, systemic, or psychiatric diseases (e.g., stroke, CVD, renal diseases, schizophrenia; see Supplementary Materials for a complete list of diseases). We thus performed the same multiple linear regression models described above separately for groups of participants with or without any of these diseases.

We furthermore assessed whether the relationship of blood pressure and mental health was dependent on the intake of antidepressants, or any other medication intake. Again, in separate multiple regression models of groups of participants taking or not taking these medications, we added SBP, HTN and number of antihypertensive medications simultaneously as predictors of depressive symptoms and well-being and corrected the models with the same covariates as described in the analyses above.

In addition, we tested whether there was a specific effect of antidepressant or antihypertensive drug classes on the relationship of blood pressure and mental health (see Supplementary Materials for list of medication classes). For these analyses, we categorised antidepressants and antihypertensives according to their mechanisms of action. In separate multiple regression models, we added SBP, HTN and a categorical variable coding for the medication classes simultaneously as predictors of depressive symptoms and well-being and corrected the models with the same covariates as described in the analyses above

All sensitivity analyses were performed on the baseline sample with measures from the initial assessment visit.

#### Exploratory analyses

##### Moderation of the BP-mental health relationship by the development of hypertension

Using moderation analysis, we explored whether the relationship between SBP and mental health was dependent on hypertension status at follow-up assessment. For this analysis, we only included participants who were <u>not</u> hypertensive at baseline, as defined by a HTN diagnosis or intake of antihypertensive medications at this timepoint. We then compared the relationship between SBP and mental health in participants who developed hypertension (i.e., received a hypertension diagnosis or started taking antihypertensives) until the follow-up assessment, with those who stayed normotensive. The moderation analysis was performed through a multiple linear regression model including depressive symptoms or well-being as outcomes, respectively, and the interaction term of SBP at initial visit and hypertension at follow-up as predictor ^42^. We also included the following covariates: age at initial visit, gender, and BMI at initial visit.

##### Associations with emotion-related brain function

Complementary to self-reports of mental health, we explored the prospective effects of elevated blood pressure on emotion-related brain function as a central nervous outcome measure of emotional reactivity. As described in the introduction, we would expect blood pressure elevations to relate to altered neural processing of affective stimuli due to baroreceptor effects. Measures of brain function included BOLD fMRI activation in significant clusters in the amygdala and whole-brain analyses (see section above *Variables*)

We used these measures as outcomes in two-sided t-tests comparing groups of participants with and without HTN, as well as in multiple linear regression models, which included SBP at follow-up (i.e., imaging visit), as well as age, gender, and BMI as covariates. In the linear regression models, we further included the interaction of SBP and HTN diagnosis at follow-up as predictor to test if the relationship between blood pressure levels and emotional reactivity differs between individuals who became hypertensive and those who did not (see also section above *Moderation of the BP-mental health relationship by development of hypertension*).

## Results

### Participants

Overall, there were data of 502,494 participants (273,378 [54.4%] women) at initial assessment visit of whom 47,933 (24,793 [51.7%] women) participants attended the follow-up visit. (Table 1) Average time between initial assessment and follow-up assessment was approximately 9 years (mean=3261 days [8.9 years], range=1400-5043 days [3.8-13.8 years]). At baseline, the median age was 58 years (range=37-73 years) and 135,745 (27%) participants reported that they had previously been diagnosed with HTN (Table 1). The final sample size for each analysis after exclusion of missing data is reported in the respective results sections below.

**Table 1.**
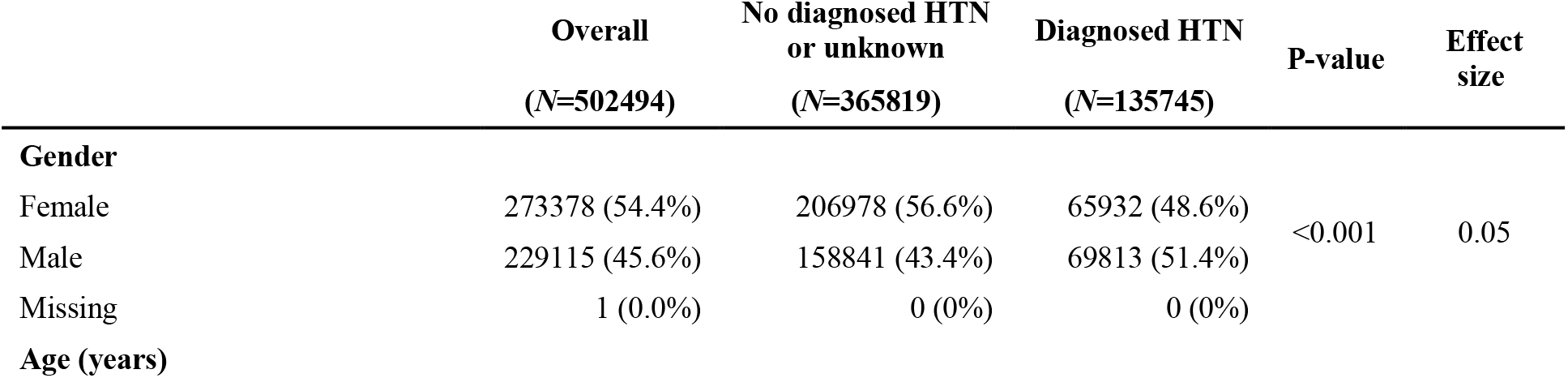
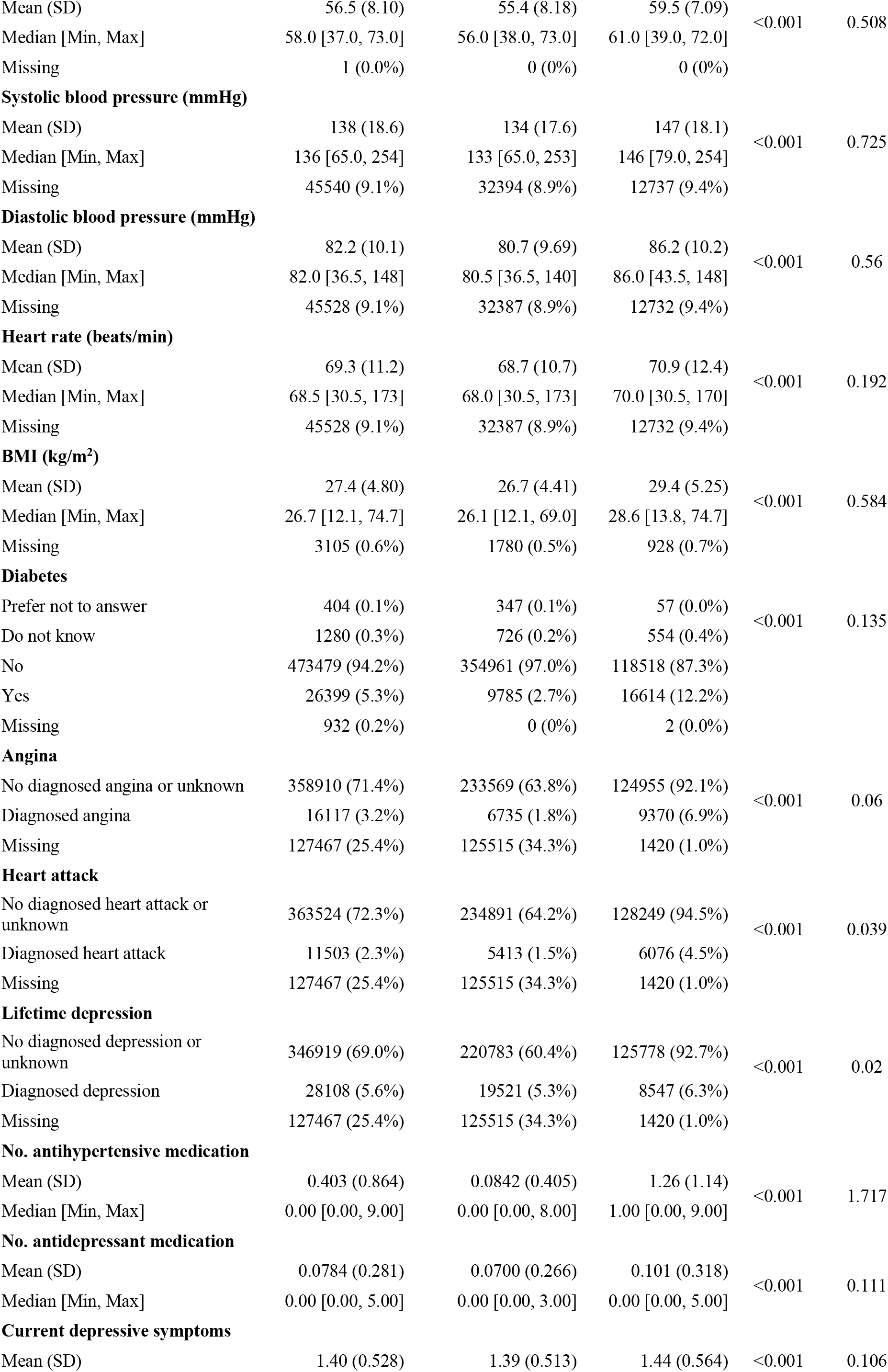
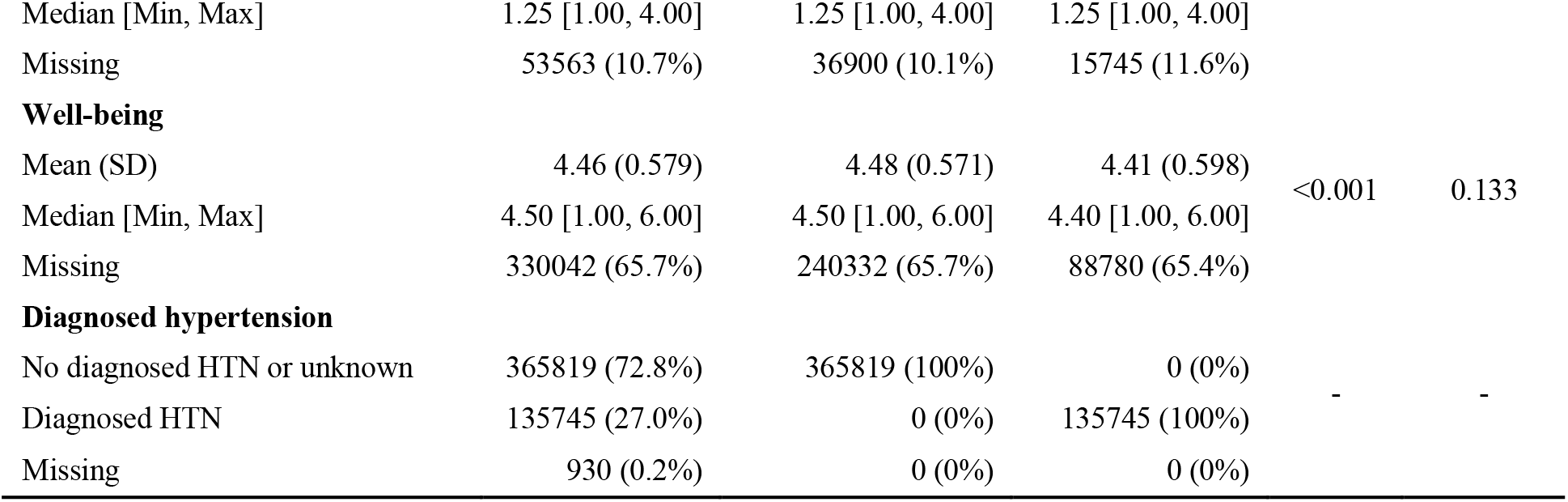
Sample characteristics at baseline assessment for total sample, as well as sub-groups with and without diagnosed hypertension (HTN). P-values and effect sizes refer to the comparison of HTN sub-groups for the respective variable. Effect sizes are specified as Cohen’s d for interval scale variables and Cramér’s V for nominal scale variables.

### Descriptive data

As shown in Table 1, participants with and without HTN diagnosis differed with respect to all descriptive variables at initial assessment. Missing data were evident in all variables of interest with varying impact (SBP 45,540 [9.1%]), HTN 930 [0.2%]), depressive symptoms 53,563 [10.7%], and well-being 385,271 [76.7%]). At follow-up assessment, missing data were evident for SBP (11,269 [23.5%]), HTN (36,264 [75.7%]), depressive symptoms (3,285 [6.9%]), and well-being (371 [0.8%]). Missing data were listwise excluded and all following analyses conducted on complete-case datasets. To account for bias due to missing data, we imputed data and conducted sensitivity analyses on the entire sample, which are reported in the Supplement. The results from imputed datasets indicated no sign of bias due to missing data and replicated our results from complete-case analyses (Supplementary Table 2).

### Outcome data

At initial assessment, self-reported current depressive symptoms resulted in a mean score of 1.40 (SD = 0.53) and well-being in a mean score of 4.46 (SD = 0.58). At follow-up, depressive symptoms were reported with mean score of 1.30 (SD = 0.44) and well-being with a mean score of 4.63 (SD = 0.54).

Among participants who completed both initial and follow-up assessments (N = 47,933), correlation analyses revealed positive associations between baseline and follow-up measures for both depressive symptoms (Pearson r = 0.529, p<0.001) and well-being (Pearson r = 0.675, p<0.001), respectively. Thus, test-retest reliability of mood assessments was high, indicating stability over assessment timepoints.

### Main results

#### Cross-sectional associations of systolic blood pressure, diagnosed hypertension, and antihypertensive medication intake with depressive symptoms and well-being

For the multiple linear regression testing cross-sectional associations of SBP, HTN, and number of antihypertensive medications with depressive symptoms 303,771 participants could be included in the model, while 129,876 could be included for the multiple linear regression model with well-being as outcome. At baseline (Figure 2), results of multiple linear regression models showed that SBP was negatively related to depressive symptoms (β = -0.063; 95% CI [-0.067, -0.060]; p<0.001) and positively associated with well-being (β = 0.057; 95% CI [0.051, 0.063]; p<0.001). Inversely, HTN was related to more depressive symptoms (β = 0.043; 95% CI [0.039, 0.047]; p<0.001) and poorer well-being (β = -0.057; 95% CI [-0.064, -0.050]; p<0.001). Restricting the analyses to participants with HTN only (N = 107,192), yielded similar associations of higher SBP with fewer depressive symptoms (β = - 0.054; 95% CI [-0.060, -0.048]; p<0.001) and greater well-being (N = 45,319), respectively (β = 0.041; 95% CI [0.032, 0.050]; p<0.001). Furthermore, our analyses yielded a small negative relationship between the number of antihypertensive medications taken and depressive symptoms (β = -0.006; 95% CI [-0.011, -0.001]; p = 0.012), and a positive trend with well-being (β = 0.007; 95% CI [0.000, 0.015]; p = 0.046). In the analyses of participants with HTN, higher numbers of antihypertensive medications were similarly associated with fewer depressive symptoms (β = -0.015; 95% CI [-0.022, -0.009]; p<0.001) and with greater well-being (β = 0.010; 95% CI [0.000, 0.020]; p = 0.043).

**Figure 2.**
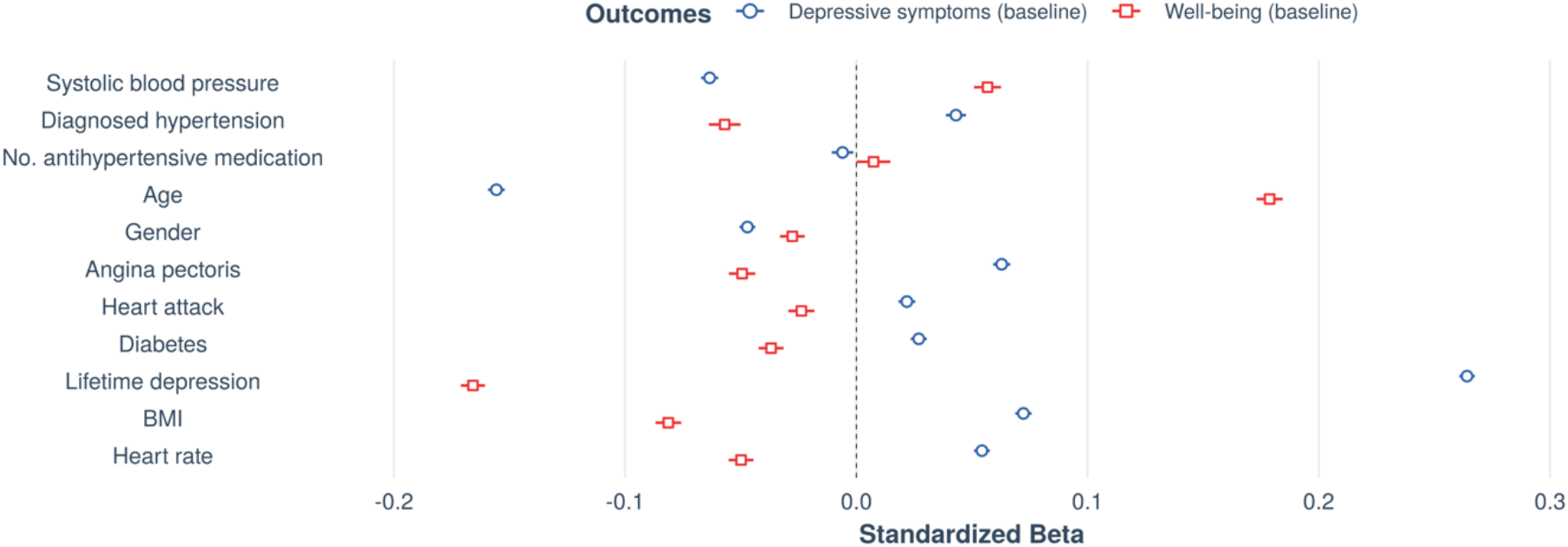
Cross-sectional associations with mental health outcomes at initial assessment. Forest plot shows standardized beta estimates and 95% confidence intervals for predictors of interest (systolic blood pressure, diagnosed hypertension (HTN), and number of antihypertensives) as well as covariates. N = 303,771 for current depressive symptoms and N = 129,876 for well-being (after exclusion of missing values).

Overall, the model for depressive symptoms including all participants yielded a model fit of adj. R^2^ = 0.129, which indicated an increase in model fit of Δadj. R^2^ = 0.004 from a null model consisting only of covariates (well-being adj. R^2^ = 0.088, Δadj. R^2^ = 0.004; HTN only: depressive symptoms Δadj. R^2^ = 0.003, well-being Δadj. R^2^ = 0.002).

#### Longitudinal associations of systolic blood pressure, diagnosed hypertension, and antihypertensive medication intake with depressive symptoms and well-being

For the multiple linear regression testing longitudinal associations of SBP, HTN, and number of antihypertensive medications with depressive symptoms 28,021 participants could be included in the model, while 29,966 could be included for the multiple linear regression model with well-being as outcome. Longitudinally (Figure 3), we found that higher SBP was related to fewer depressive symptoms at follow-up assessment (β = -0.042; 95% CI [-0.055, -0.029]; p<0.001) and to higher follow-up well-being scores (β = 0.033; 95% CI [0.020, 0.046]; p<0.001). Similar to the cross-sectional results, baseline HTN was associated with more depressive symptoms approximately 10 years later (β = 0.029; 95% CI [0.014, 0.044]; p<0.001). HTN was also significantly related to lower well-being scores at follow-up (β = -0.032; 95% CI [-0.047, -0.017]; p<0.001). Number of antihypertensive medications at baseline was not a significant predictor in any longitudinal model (depressive symptoms: β = -0.006; 95% CI [-0.022, 0.009]; p=0.418; well-being: β = -0.004; 95% CI [-0.019, 0.011]; p=0.620). The model fit for the prediction of depressive symptoms increased from a null model consisting only of covariates by Δadj. R^2^ = 0.001 (adj. R^2^ = 0.092; well-being adj. R^2^ = 0.055, Δadj. R^2^ = 0.001).

**Figure 3.**
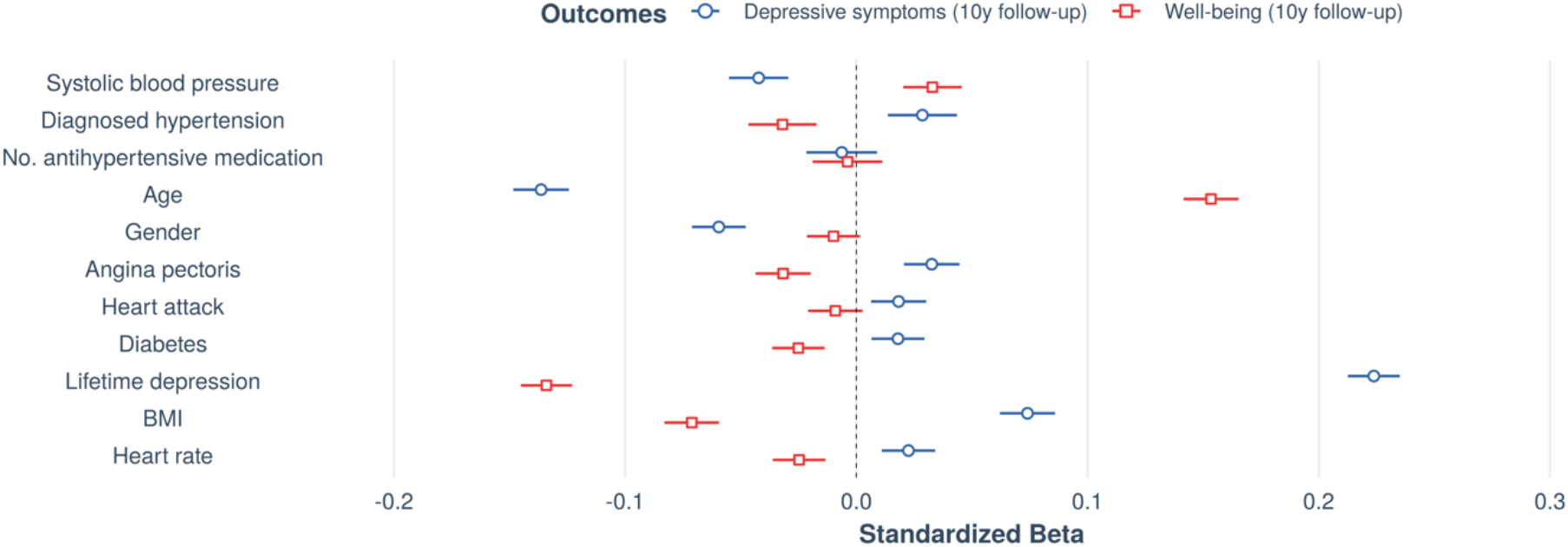
Longitudinal associations with mental health outcomes at follow-up assessment. Forest plot shows standardized beta estimates and 95% confidence intervals for predictors of interest at baseline (systolic blood pressure, diagnosed hypertension (HTN), and number of antihypertensives) as well as covariates. N = 28,021 for current depressive symptoms and N = 29,966 for well-being (after exclusion of missing values).

#### Additional and sensitivity analyses

We performed several additional analyses to test the robustness of these results. Additional analyses included i) using the PHQ-9 questionnaire as a validated instrument to assess current depressive symptoms, ii) additional relevant covariates, such as socioeconomic status, insomnia, racial/ethnic background, insomnia, etc., iii) using hospital inpatient data for diagnoses of HTN and depression, iv) assessment of potential survival bias and v) exploration of potential unmeasured confounding effects with E-values. Moreover, sensitivity analyses were performed to test whether the above reported results were dependent on vi) the presence or absence of previous diagnosis of depression or any other severe disease that might affect BP (list of diseases in Supplementary Materials); vii) the intake of antidepressants or any other medication intake; viii) a specific effect of certain antidepressant or antihypertensive drug classes.

The results of all additional and sensitivity analyses are reported in the Supplementary Materials. In sum, the results were overall robust and consistent: independent of any potential confounders including medication intake, there was an effect of higher SBP on fewer depressive symptoms and higher well-being, as well as a negative effect of HTN diagnosis on mental health.

### Exploratory analyses

#### Moderation of the BP-mental health relationship by development of hypertension

Next, we explored if the relationship between mental health and SBP was moderated by hypertension status at follow-up assessment. First, we excluded participants who were hypertensive at the initial assessment (defined as HTN diagnosis or intake of antihypertensives). This resulted in 315,582 people who could be considered non-hypertensive at initial assessment and who were included in the analysis (of those, N = 25,991 had data available to define hypertension at follow-up). Across all participants, SBP increased from initial assessment to follow-up (Figure 4A, mean increase = 3.820 mmHg; *t* = 38.996; degrees of freedom = 25578; 95% CI [3.628, 4.012]; *p*<0.001). People who later developed HTN already had higher SBP levels at initial assessment compared to people who stayed normotensive (Figure 4B, HTN: mean baseline SBP (SD) = 148 (18.7) mmHg; no HTN: mean baseline SBP (SD) = 130 (15.3) mmHg). Notably, in unadjusted models, there were no significant group differences in mental health at initial assessment between people who developed HTN and those who stayed normotensive (Figure 4C, HTN: mean depressive symptoms = 1.333; no HTN: mean depressive symptoms = 1.327; *t* = -0.738; degrees of freedom = 4585.9; 95% CI [-0.023, 0.010]; *p*=0.464; Figure 4D, HTN: mean well-being = 4.536; no HTN: mean well-being = 4.547; *t* = 0.662; degrees of freedom = 1583.8; 95% CI [-0.022, 0.044]; *p*=0.508). However, in the fully adjusted regression model, we observed a main effect of later HTN on baseline mental health (Figure 4C, depressive symptoms: β = 0.060; 95% CI [0.045, 0.076]; *p*<0.001; Figure 4D, well-being: β = -0.043; 95% CI [-0.068, -0.017]; *p*<0.001), suggesting that when adjusting for SBP levels, people who later developed HTN had lower mood (i.e. more depressive symptoms and lower well-being) already at initial assessment compared to people without HTN. The regression model further yielded a significant interaction of SBP at initial assessment and HTN at follow-up with depressive symptoms at initial assessment, showing that the association between SBP and depressive symptoms was moderated by hypertension status at follow-up (Figure 4E, β = -0.014; 95% CI [-0.026, -0.003]; p=0.015). Thus, the slope of the relationship between higher SBP and fewer depressive symptoms was steeper in those participants who developed HTN approximately 10 years later. At follow-up, however, HTN status was not a significant moderator for the association of SBP and depressive symptoms (β = -0.005; 95% CI [-0.016, 0.007]; *p*=0.419). Similarly, a trend of a moderation by HTN status was observed for the association between well-being and SBP at initial assessment, but this effect was not significant (Figure 4F, β = 0.017; 95% CI [-0.001, 0.036]; p=0.070). Neither was there a moderation by HTN status for the association between well-being at follow-up and SBP at follow-up (β = 0.002; 95% CI [-0.010, 0.013]; p=0.770).

**Figure 4.**
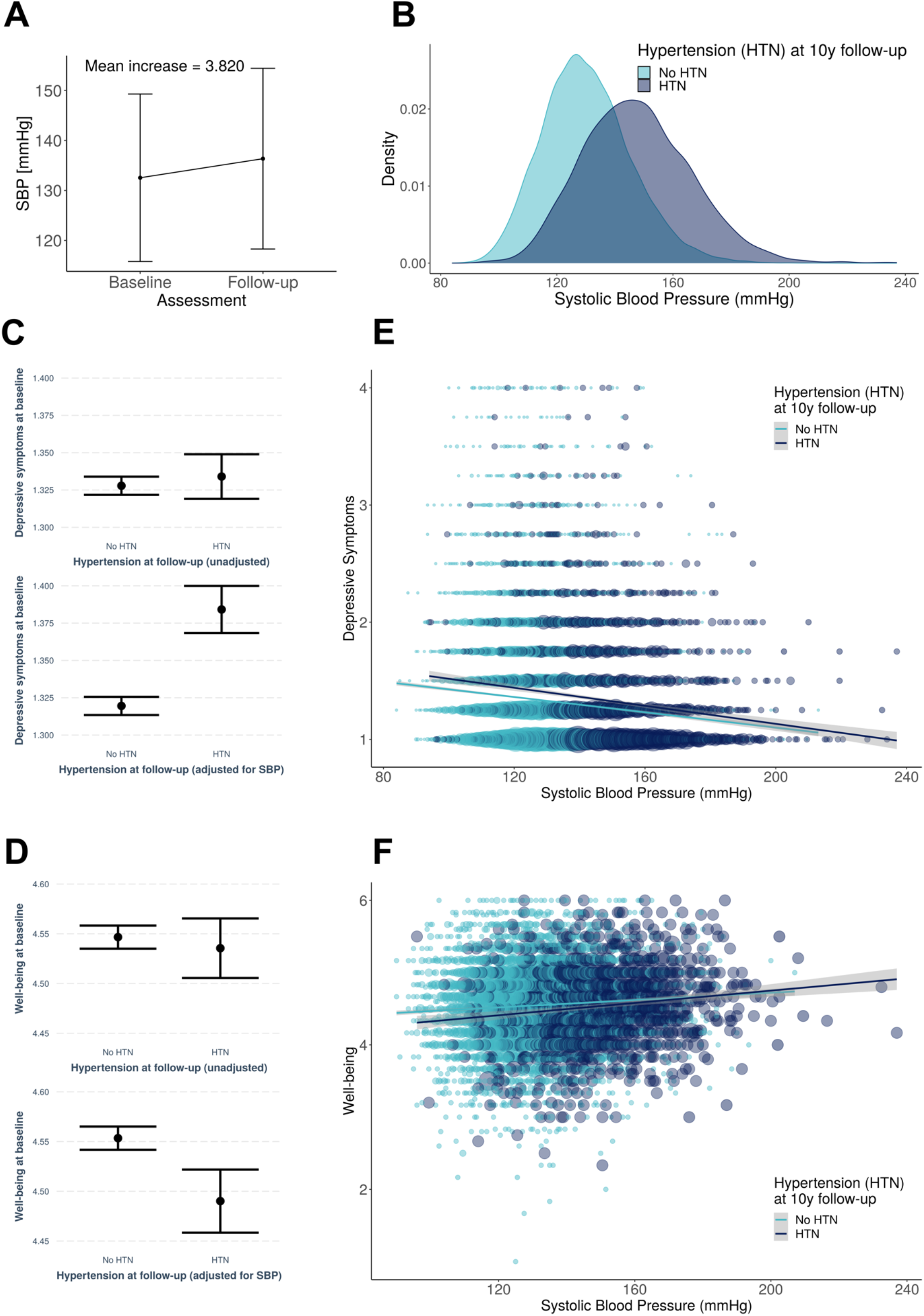
Association between mental health and systolic blood pressure at initial assessment moderated by hypertension status at follow-up (i.e. approximately 10 years later). A: Across all participants (excluding those with HTN and use of antihypertensives), systolic blood pressure increased from baseline to follow-up. B: People who developed hypertension until follow-up (i.e. received a hypertension diagnosis or started taking antihypertensives) until the follow-up assessment (dark blue colour) already showed higher systolic blood pressure levels at initial assessment, despite not having been diagnosed at this timepoint, yet. C: No significant difference between groups in depressive symptoms at baseline (top), but when controlling for SBP, HTN developers showed more depressive symptoms (bottom). D: No significant difference between groups in well-being at baseline (top), but when controlling for SBP, HTN developers showed lower well-being (bottom). E: Participants who developed hypertension (dark blue colour) had a steeper negative slope for the relationship between blood pressure and depressive symptoms than those participants who stayed non-hypertensive (light blue colour). F: Similar trend for well-being in the expected opposite direction.

We replicated these findings using SBP > 140 mmHg as an additional criterion to define hypertension. This resulted in a sample where the two groups had SBP levels in the normotensive range at baseline. The results within this sample remained virtually unchanged and are reported in detail in the Supplementary Results.

#### Associations with emotion-related brain function

Finally, to explore a central nervous representation of emotional reactivity, we assessed how hypertension status relates to emotional processing in the brain using fMRI activity assessed during the Hariri task. BOLD fMRI activity to emotional faces was lower in the amygdala (HTN = 1.134; no HTN = 1.249; t = 9.797; degrees of freedom = 15,129; 95% CI [0.091, 0.137]; p<0.001) and in significant regions resulting from whole-brain analyses (HTN = 2.322; no HTN = 2.592; t = 14.528; degrees of freedom = 14,888; 95% CI [0.233, 0.306]; p<0.001) in people with HTN compared to normotensives (Figure 5 left panels). We also observed a negative association between lower BOLD reactivity to emotional faces and higher SBP across participants, suggesting a continuously negative effect of SBP on BOLD (amygdala: β = -0.041 95% CI [-0.054, -0.028]; p<0.001; whole-brain: β = -0.032 95% CI [-0.045, -0.019]; p<0.001, Figure 5 middle panels). Additionally, an interaction of HTN and SBP suggested that the negative association between SBP and whole-brain BOLD activity was less pronounced in people with HTN (whole-brain: β = 0.015 95% CI [0.003, 0.028]; p=0.014; trend in amygdala: β = 0.024 95% CI [-0.001, 0.024]; p=0.063; Figure 5 right panels). We repeated the same analysis using the blood pressure measurement at initial assessment which was acquired ∼10 years prior to neuroimaging and thus not directly related to blood flow and volume dependent effects during BOLD fMRI, Interestingly, already the baseline SBP (i.e., prior to any HTN diagnosis) was negatively correlated with BOLD reactivity to emotional faces. In addition, there was an interaction of baseline SBP and later HTN on emotion-related brain activity, specifically in amygdala regions (β = 0.017; 95% CI [0.004, 0.030]; p=0.009), suggesting that the negative correlation between SBP at baseline and emotion-related amygdala activity was dampened in participants who developed hypertension. The interaction between baseline SBP and BOLD activity was not significant in the whole-brain analysis (β = 0.008; 95% CI [-0.005, 0.021]; p=0.213).

**Figure 5.**
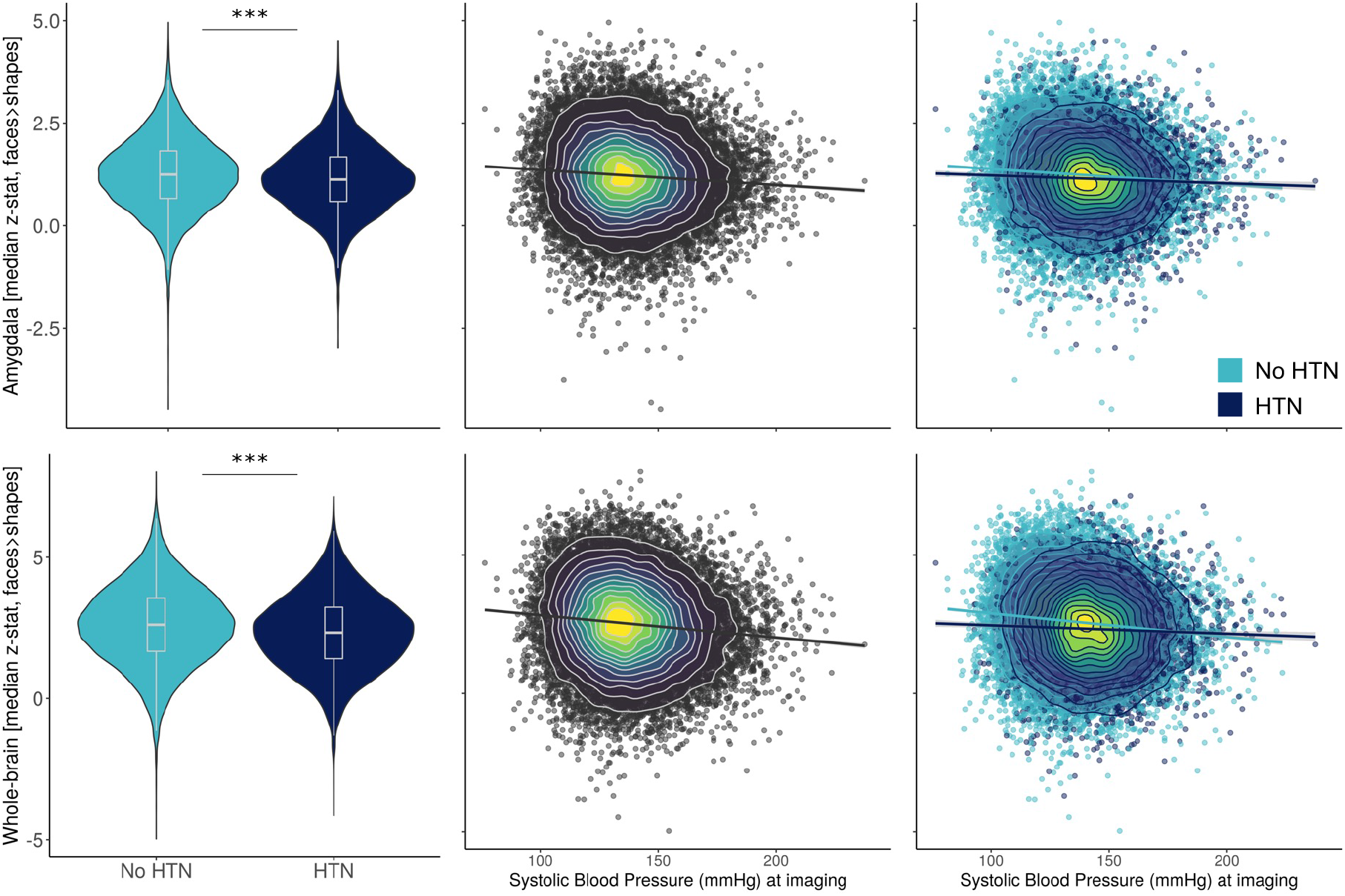
Association between systolic blood pressure and emotion-related brain function (assessed at follow-up approximately 10 years later). Left column (violin plots) shows group differences in Hariri task activity by hypertension status (HTN) at follow-up/imaging visit. Middle column shows negative correlations between blood pressure and BOLD fMRI activation in the faces>shapes contrast of the Hariri task in amygdala mask and whole-brain mask. The colour gradient represents the density of data points. Right column shows the same, but grouped by HTN at follow-up. Dark blue colours represent people who became hypertensive from baseline to follow-up. Light blue colours represent participants who stayed normotensive. The negative correlation between systolic blood pressure and emotion-related activity was flattened in participants who had developed hypertension.

## Discussion

In this study, we confirmed two seemingly contradictory associations of high blood pressure with mental health: (i) Higher SBP was associated with fewer depressive symptoms and greater well-being at the initial exam as well as at the 5-year mental health online follow-up and the 10-year follow-up including imaging, whereas (ii) the presence of a HTN diagnosis was associated with greater depressive symptoms and lower well-being. Given the well-powered cohort of the UK Biobank, we were able to perform sensitivity analyses which confirmed that the observed associations were robust to influences of medication intake (e.g., antihypertensives, antidepressants) and chronic diseases (e.g., CVD, clinical depression). Strikingly, we furthermore found that (iii) already at the initial exam, among normotensive individuals, mental health was negatively affected (more depressive symptoms, lower well-being) by the *later* HTN status at 10y-follow-up. Also, (iv) the relationship between blood pressure and mental health – at the initial exam – was moderated by later HTN, such that the negative association between blood pressure and depressive symptoms was stronger in those participants who later developed HTN. Finally, (v) our results from task-based functional brain imaging provide further support for an impact of blood pressure and HTN on central processing of emotions: We demonstrate a negative relationship between SBP - at baseline (∼10 years prior to fMRI) and at the time point of imaging - and the BOLD fMRI response to aversive emotional stimuli. This relationship was again moderated by HTN status at follow-up, such that people who developed HTN showed overall lower responses to aversive stimuli and a flattened (negative) relationship between SBP and brain responses. Taken together, our results support the notion that the interrelation between blood pressure and mental health may be involved in the development of high blood pressure with potential implications for developing new preventive and therapeutic approaches for essential hypertension.

We preregistered and confirmed a conceptional replication that higher SBP related to better mood ratings: Our findings are consistent with studies reporting positive effects of elevated blood pressure on mental health, including decreased depressive symptoms ^16–18^, better quality of life ^12,18^, and reduced self-reported stress ^13–15^. We extend these findings, which were based on either cross-sectional designs or shorter follow-up periods (1-5 years), by demonstrating that baseline SBP remained a significant predictor of mental health up to 10 years later. The positive effects of high blood pressure on mood might be related to research findings showing that higher blood pressure diminishes emotional experience in experimental manipulations ^43–45^. It is also well established that elevated blood pressure robustly reduces the perception of physical pain ^21,25,46,47^, but also social pain ^48^. It has therefore been hypothesised that elevated blood pressure relates to a generalised attenuation of emotional valence processing ^25,45^. Our fMRI findings are consistent with this notion, suggesting an impact of blood pressure on the processing of (negative) emotional faces, even at a follow-up after 10 years. Importantly, while affective attenuation might relate to coping mechanisms to ‘lift mood’ in stressful situations, it could reinforce staying in potentially harmful circumstances over prolonged periods of time. This may lead to further blood pressure increases and eventually to HTN (see below) ^20,25,27–29^.

Given these consistent findings of a positive association between blood pressure and mood, it seems paradoxical that we found a different pattern for diagnosed HTN; albeit again in line with previous studies in which the presence of vascular risk factors or manifest CVD has been associated with increased depressive symptoms ^49,50^ and decreased well-being ^51^. Several potential explanations have been put forward: Biological explanations build on well-known pathophysiological consequences of chronic blood pressure elevations (e.g., atherosclerosis, small vessel disease, etc.) leading to ischaemic brain damage indicated by white matter lesions, microinfarcts, and cerebral micro-haemorrhages ^52^. White matter lesions, in particular, have been linked to the occurrence of depression with a vascular component ^53,54^. Systemic mechanisms resulting from unfavourable metabolic alterations, which are common in people with HTN, and an unhealthy lifestyle (e.g. smoking, physical inactivity, unbalanced diet, etc.) have also been linked to depression via metabolic, immuno-inflammatory and autonomic pathways (reviewed in ^55^). Psychological explanations, on the other hand, emphasise that individuals receiving a diagnosis of HTN are often confronted with a sudden awareness of a chronic illness that requires medical attention and change of lifestyle. The negative psychological consequences of such a ‘labelling’ effect could underlie opposing effects of elevated blood pressure and HTN diagnosis on mental health ^56,57^.

Based on our data, we cannot exclude contributions of the factors discussed above, however, a major new finding of our study (i.e., that an impact of HTN on mood is already present before the diagnosis), is difficult to reconcile with these explanations: While in our analysis, we observed – as expected – that, at the initial visit, those participants who later developed HTN already had higher blood pressure than those who stayed normotensive,; in the fully adjusted model, i.e., correcting for the differences in blood pressure, the negative impact of (later) HTN on mental health was already significantly present *before* the HTN diagnosis. We found this in two analyses, one in which HTN was defined by previous HTN diagnosis and intake of medication, and one in which SBP>140 mmHg was also used as criterion. In both analyses – when unadjusted for blood pressure – there were no significant differences in mental health at either visit while the difference was highly significant when adjusted for blood pressure. Interestingly, we also noted in both analyses that the two groups differed regarding the overall ‘mood-lifting effect’ of higher blood pressure at the initial visit, such that this effect was more pronounced in the group of those participants who developed HTN later. Thus, it seems that in the HTN-developing group, the relationship between blood pressure and mental health was both shifted in magnitude and had a different slope. This finding cannot be explained by the ‘labelling’ effect and is also unlikely to be related to vascular damage, such as white matter lesions, as these occur only after long-lasting blood pressure elevations.

Obvious candidates for explaining effects of blood pressure on mental health are regulatory circuits linking arterial blood pressure to central processing in the brain. While the causal, and likely multifactorial, pathways between blood pressure and mental health are not fully understood, a shared mechanism between subjective experience, emotional processing and pain involves the regulatory baroreflex system. Baroreceptors, stretch sensitive receptors located in the aortic arch and the carotid artery sinus, are the peripheral sensors of blood pressure ^58–60^. During each heartbeat, baroreceptors are activated during systole and become less active during diastole. They are known to relay phasic and tonic information about blood pressure via the vagal and glossopharyngeal nerves to brain stem nuclei which orchestrate adjustments of blood pressure and heart frequency via the parasympathetic and sympathetic nervous system ^58–60^. Importantly, in addition to their role in adjusting blood pressure and heart frequency, baroreceptor activation has also been shown to influence emotional and pain processing (reviewed by Suarez-Roca et al., 2021), thereby mediating behavioural and central effects of blood pressure modulations. Direct evidence comes from animal studies, in which for example, pain-relieving effects of blood pressure elevations were abolished by baroreceptor denervation ^20,27^ and from studies in humans in whom local baroreceptor stimulation modulated pain perception ^21,24,25,27^. Further evidence has been provided by numerous studies showing different processing of pain, emotion, and sensory stimuli in systole versus diastole ^22,23,26,61^. Importantly, it is also well established that the development of HTN is characterised by a progressive desensitisation of baroreceptors and altered sensory processing ^27,62^. It, therefore, seems highly plausible that (relatively reduced) baroreceptor signalling might also underlie the observed altered relationship between blood pressure and mental health in hypertensive people. Our results furthermore indicate that the altered relationship between blood pressure and mental health may already be present years before the diagnosis of HTN. In a similar vein, our fMRI findings show an altered relationship between blood pressure and BOLD activation to negative facial expressions in people with HTN, consistent with adjusting central processing of emotions as a response to baroreceptor desensitisation. In sum, there is evidence that baroreceptor signalling can underlie the effect of higher blood pressure on mental health.

With regard to the development of blood pressure increases over the life course and eventually the pathophysiology of arterial HTN, our findings are consistent with the notion of feedback loops wherein arousing emotional stimuli and stressors elevate blood pressure, which in turn activates baroreceptor pathways that induce analgesia and decrease the perceived affective magnitude of a stressor ^29^. While psychosocial stress is increasingly accepted as a risk factor for the development of hypertension ^63^, blood pressure adjustments – via baroreceptor signalling - may link stress to a rewarding mechanism decreasing perceived stress. Reinforcement by repeated stress exposure may eventually lead (or contribute) to increases in blood pressure and the development of essential HTN ^20,25,27–29^. Our data further emphasise inter-individual differences: Those individuals who later developed HTN on average showed lower mental health scores when adjusting for SBP. In addition, our moderation analysis yielded that the development of HTN was associated with a stronger negative correlation between mental health and blood pressure at baseline, years before the HTN diagnosis. While the observed *inter*-individual differences cannot be readily interpreted as indicative of *intra*-individual mechanisms, it is nevertheless tempting to speculate that people at higher risk of developing HTN require higher blood pressure levels to sustain the same mental health outcomes. These individuals may find themselves on a relatively steeper trajectory towards HTN due to the stronger ‘mood-lifting effect’ of blood pressure increases. Taken together we propose that (i) feedback loops between blood pressure and rewarding emotional processing during periods of stress may play a role in the pathophysiology of blood pressure increases and HTN and that (ii) alterations of these feedback loops characterised by a shifted blood pressure - mental health relationship may increase HTN risk in affected individuals. Based on our data, we cannot differentiate between potential reasons for the altered blood pressure-mental health relationship in the mostly middle-aged participants, which may include genetics, life-style factors, such as nutrition or the environment, previous exposure to acute and chronic stress or other factors. Clearly, there is a need for prospective longitudinal studies clarifying this issue.

Beyond effects of blood pressure and diagnosed HTN, other factors such as medication and previous diseases can influence mental health and thus be confounders in our analyses. For example, Hermann-Lingen et al. ^18^ reported lower physical well-being in participants on antihypertensive medication. Boal et al. ^64^ reported differential effects of antihypertensive medication on risk of hospital admissions for mood disorders. Additionally, intake of antidepressant drugs has been previously related to elevated blood pressure ^16^. To account for these potentially confounding factors, we investigated effects of the presence or absence of a lifetime major depression diagnosis, other forms of chronic illness as well as intake of (antihypertensive) medications in sensitivity analyses. Importantly, the findings of our study were robust and consistent: Independent of any potential confounders including medication intake, there was a ‘mood-lifting’ effect of higher SBP on depressive symptoms and well-being, as well as a negative effect of HTN diagnosis on mood.

The results reported here are contingent on several limitations. We used UK Biobank data which is not representative of the middle-aged and older UK population ^65^. The sample, particularly the neuroimaging sub-sample, displays the ‘healthy volunteer’ effect, which describes that UK Biobank participants are considered to be more health-conscious, self-reported fewer health conditions and show lower rates of all-cause mortality and total cancer incidence compared to the general population ^65^. Yet, associations between risk factors and disease outcomes in UK Biobank have been reported to be generalisable despite the ‘healthy volunteer’ effect ^65,66^.

Similar to previous studies, we used self-reports of HTN and antihypertensive medications. Self-reported HTN can underestimate the true underlying prevalence ^67^ and may influence subjective health itself ^57^. In addition, participants’ self-reports of prescribed antihypertensive medications might overestimate the actual number of medications taken due to poor adherence ^68^. In addition to self-report measures, we included direct standardised blood pressure recordings, as well as Likert-scale measures of depressive symptoms and well-being, which enabled us to parametrically model these exposures and outcomes in linear regression models. The mental health assessments were, however, not designed for psychiatric diagnostics, which leaves the question open whether the observed effects manifest in the sub-clinical and/or clinical range of psychiatric symptoms.

A recent study also showed that averaging blood pressure values from the first and second reading, as we did here, might underestimate the true prevalence of HTN ^69^. While we have not used the blood pressure readings to define HTN in our study, we acknowledge that the procedure of blood pressure readings may have an undetected effect on our results. Yet, our results converge also when using HTN diagnosis from self-reports and hospital records, which strengthens the overall confidence in the robustness of our findings.

Conceptually, one may question the strict dichotomy between HTN versus no HTN, particularly as blood pressure thresholds for HTN diagnosis have shifted towards lower values in the last decades. However, given the diagnostic criteria for HTN at the time of the study, we assume them to be followed by most physicians in clinical care. Thus, we consider the self-reported HTN status to be a reasonable definition with predictive value for clinical outcomes in an epidemiological study ^68^.

Based on our preregistered hypotheses, we only tested linear associations between blood pressure and mental health. While Montano ^16^ showed that non-linear models testing cross-sectional blood pressure-mood associations do not outperform linear models, longitudinal trajectories of both blood pressure, mental health and their interaction over the lifespan are plausible and may reveal diverging patterns. Furthermore, observed effect sizes were small and currently clearly no conclusion can be made from our data on individual patient care. Given the known effects of blood pressure on emotional processing, we speculate that – despite the small inter-individual effect sizes in our study – more pronounced intra-individual effects might exist. Our study may stimulate future work testing the hypothesis that blood pressure variations and associated mental health need to be taken into account, also in the individual management of people at risk for HTN. Considering the high prevalence of HTN and its treatment in the general population, as well as rising numbers of sub-clinically elevated blood pressure, small effect sizes may be epidemiologically relevant.

Our fMRI findings may be confounded by alterations in neurovascular coupling with HTN ^70^. While this should be largely accounted for by reporting the BOLD activation *contrast* between two different types of stimuli (emotional faces versus shapes), we cannot exclude a remaining impact of impaired neurovascular coupling. Since the Hariri-task was limited to faces with negative emotions, future studies may add information about the impact of blood pressure on neural processing of positive emotions.

Importantly, our results are not ideal to draw firm conclusions about causality and directionality of the associations between blood pressure, HTN and mental health, with particularly the differentiation between effects of blood pressure *per se* and HTN remaining complex. Future longitudinal studies should therefore include earlier baseline assessments, repeated and/or continuous blood pressure monitoring over long time periods combined with frequent assessments of mental health and neuroimaging. Finally, randomised controlled trials targeted at assessing the bi-directional relationships of blood pressure and mental health will provide strong designs to elucidate these effects.

In summary, in a large British population sample of generally healthy middle-aged and older individuals, we found a relationship of elevated blood pressure with fewer depressive symptoms and greater well-being extending to a follow-up period up to around 10 years. We found the opposite effect for diagnosed HTN – already years before the timepoint of diagnosis. Participants who were normotensive at baseline and later developed HTN showed alterations in the blood pressure-mental health relationship already at baseline. An additional novelty of our study lies in the use of fMRI analyses, which suggested an impact of blood pressure levels and HTN development on the neural processing of emotional stimuli. The results were overall robust to bias of medications, chronic illness, survival, social factors and unmeasured confounds. While the observed effects are small and results from this observational study may not be directly applicable to clinical outcomes, our study provides a novel perspective on how the interrelation of sub-clinical mental health and blood pressure might be involved in blood pressure increases during ageing and development of HTN that could have implications for developing new preventive and therapeutic approaches.

## Data Availability

All data used in this study is available through the public resource of the UK Biobank (http://www.ukbiobank.ac.uk/). The authors' access to the UK Biobank Resource was granted under Application Number 37721.

## Code Availability

To allow for replication studies and reproducibility of our results, the analysis code can be found in the Open Science Framework repository of this study (https://osf.io/638jg).

## Acknowledgements

This research has been conducted using the UK Biobank Resource under Application Number 37721. We would like to sincerely thank all participants and staff who made it possible that this rich resource is available to the scientific community.

## Author contributions

L.S., M.B., D.K., M.U. and A.V. conceptualized the study. L.S., M.B., D.K., M.U. and A.V. designed and developed the methodologies. L.S. analysed, visualised, and curated the data, as well as managed the research project. L.L. curated the medication data. L.S. and A.V. wrote the initial draft of the manuscript. A.V. and S.L.V. supervised the research. All authors critically reviewed and edited the manuscript and contributed to the final version of the paper.

## Competing interests

The authors declare no competing interests.

## Notes

### Competing Interest Statement

The authors have declared no competing interest.

### Funding Statement

This study did not receive any external funding.

### Author Declarations

All data used in this study is available through the public resource of the UK Biobank. Ethical approval for the UK Biobank has been granted by the North West Multi-centre Research Ethics Committee (MREC). For details please see: https://www.ukbiobank.ac.uk/learn-more-about-uk-biobank/about-us/ethics

