## Supplementary Materials for "Mental health, blood pressure and the development of hypertension"

H. Lina Schaare[1,7,8,11], Maria Blöchl[1,2], Deniz Kumral[1,9,10], Marie Uhlig[1], Lorenz Lemcke[4], Sofie L. Valk[7,8,11], Arno Villringer[1,3,5,6]

Affiliations

*1 Department of Neurology, Max Planck Institute for Human Cognitive and Brain Sciences, Leipzig, Germany*
*2 Institute for Psychology, Leipzig University, Leipzig, Germany*
*3 MindBrainBody Institute, Berlin School of Mind and Brain, Berlin, Germany*
*4 Nuclear Magnetic Resonance Unit, Max Planck Institute for Human Cognitive and Brain Sciences, Leipzig, Germany*
*5 Clinic of Cognitive Neurology, Leipzig University, Leipzig, Germany*
*6 Charité University Medicine Berlin, Berlin, Germany*
*7 Otto-Hahn-Group Cognitive Neurogenetics, Max Planck Institute for Human Cognitive and Brain Sciences, Leipzig, Germany*
*8 Institute of Neuroscience and Medicine (INM-7: Brain and Behaviour), Research Centre Jülich, Germany*
*9 Institute of Psychology, Neuropsychology, University of Freiburg, Freiburg, Germany*
*10 Institute of Psychology, Department of Clinical Psychology and Psychotherapy, University of Freiburg, Freiburg, Germany*
*11 Institute of Systems Neuroscience, Medical Faculty, Heinrich Heine University Düsseldorf, Düsseldorf, Germany*

Corresponding author: H. Lina Schaare
Max Planck Institute for Human Cognitive and Brain Sciences
Stephanstr. 1A, 04103 Leipzig, Germany
+4934199402220


#### Contents

Mental health, blood pressure and the development of hypertension ... 1
Contents ... 2
Supplementary Methods ... 3
UK Biobank fields used in this study ... 3
Multiple Imputation of missing data ... 5
Diagnoses and details on medication intake for sensitivity analyses ... 6
Medication intake ... 8
Antihypertensive drug classes ... 9
Antidepressant drug classes ... 10
Supplementary Results ... 11
PHQ-9 results ... 11
Modelling of additional relevant variables ... 11
Analyses with hospital records (HES) ... 12
Assessment of survival bias ... 14
Moderation analysis of BP-mental health relationship with SBP as additional criterion to define hypertension status ... 16
Inclusion/exclusion of previous depression and other BP-altering diseases ... 17
Medication effects ... 18
Exploration of unmeasured confounding bias with the E-value ... 20
References ... 21

### Supplementary Methods

#### *UK Biobank fields used in this study*

*Depressive symptoms*

During each assessment centre visit (at 1. initial assessment visit (2006-2010), 2. imaging visit (2014+)), questions were asked about the frequency of depressive symptoms in the last two weeks (i.e. depressed mood [http://biobank.ctsu.ox.ac.uk/crystal/field.cgi?id=2050], unenthusiasm [http://biobank.ctsu.ox.ac.uk/crystal/field.cgi?id=2060], tenseness [http://biobank.ctsu.ox.ac.uk/crystal/field.cgi?id=2070], tiredness [http://biobank.ctsu.ox.ac.uk/crystal/field.cgi?id=2080]) using a 4-point Likert scale ranging from 0 ("not at all") to 3 ("nearly every day"). For this study, current depressive symptom frequency scores were summarized as mean scores.

We also preregistered analyses using the Patient Health Questionnaire 9-question version (PHQ-9) (Kroenke et al., 2001) from the online mental health follow-up assessment which a sub-sample of the whole UK Biobank cohort received (Davis et al., 2020). In the online PHQ-9 questionnaire, severity of current depressive symptoms in the last two weeks was assessed. Participants indicated presence of recent depressive symptoms on a 4-point Likert scale ranging from 0 ("not at all") to 3 ("nearly every day") [PHQ-9 items: http://biobank.ctsu.ox.ac.uk/crystal/label.cgi?id=138]. Since UK Biobank applied a different coding scheme, the items were recoded to match the original coding as described above. PHQ-9 symptom scores were summarized as a sum score.

*Well-being*

Seven questions addressing different aspects of participants' well-being were included at three instances of UK Biobank data acquisition (at 1. initial assessment visit (2006-2010), 2. first repeat assessment visit (2012-2013), 3. imaging visit (2014+)). The questions included happiness [http://biobank.ctsu.ox.ac.uk/crystal/field.cgi?id=4526], health satisfaction [http://biobank.ctsu.ox.ac.uk/crystal/field.cgi?id=4548], work satisfaction [http://biobank.ctsu.ox.ac.uk/crystal/field.cgi?id=4537], family satisfaction [http://biobank.ctsu.ox.ac.uk/crystal/field.cgi?id=4559], friendship satisfaction [http://biobank.ctsu.ox.ac.uk/crystal/field.cgi?id=4570], and financial situation satisfaction [http://biobank.ctsu.ox.ac.uk/crystal/field.cgi?id=4581]. Participants were asked to respond to the questions on a 6-point Likert scale ranging from 1 ("extremely happy") to 6 ("extremely unhappy"). For this study, well-being scores were recoded and summarized as a mean score with higher scores representing greater happiness and satisfaction.

*Systolic blood pressure*

Systolic and diastolic blood pressure readings were taken from all UK Biobank participants. At each assessment visit (1. at baseline assessment (2006-2010), 2. first repeat assessment visit (2012-2013), 3. imaging visit (2014+)), two readings were recorded with an automated blood pressure monitor (sphygmomanometer) a few moments apart during seated resting periods. We used the mean of the two readings to derive systolic (SBP) values per participant at each visit. [https://biobank.ctsu.ox.ac.uk/crystal/field.cgi?id=4080]

*Hypertension diagnosis*

Self-reports at each assessment visit were used for diagnosis of HTN. Participants were asked whether a doctor has ever told them that they have had high blood pressure (touchscreen question, [https://biobank.ctsu.ox.ac.uk/crystal/field.cgi?id=6150]).

*Number of antihypertensive medications*

In a nurse interview at each assessment visit, participants reported all medications that they were currently taking [https://biobank.ctsu.ox.ac.uk/crystal/field.cgi?id=20003]. A physician from our team (LL) examined these medication lists and identified all antihypertensive drugs. We subsequently coded these antihypertensives and calculated a sum score for each participant.

*Confounding variables*

In addition, we considered confounding effects of the following variables:

age [http://biobank.ctsu.ox.ac.uk/crystal/field.cgi?id=21003], gender [http://biobank.ctsu.ox.ac.uk/crystal/field.cgi?id=31], diabetes status [http://biobank.ctsu.ox.ac.uk/crystal/field.cgi?id=2443], history of diagnosed angina [http://biobank.ctsu.ox.ac.uk/crystal/field.cgi?id=20002], history of diagnosed myocardial infarction [http://biobank.ctsu.ox.ac.uk/crystal/field.cgi?id=20002], history of diagnosed depression [http://biobank.ctsu.ox.ac.uk/crystal/field.cgi?id=20002], body-mass index [http://biobank.ctsu.ox.ac.uk/crystal/field.cgi?id=21001], resting heart rate [http://biobank.ctsu.ox.ac.uk/crystal/field.cgi?id=102], history of diagnosed depression [http://biobank.ctsu.ox.ac.uk/crystal/field.cgi?id=20002], current intake of antihypertensive and antidepressive medication [http://biobank.ctsu.ox.ac.uk/crystal/field.cgi?id=20003], history of severe disease [http://biobank.ctsu.ox.ac.uk/crystal/field.cgi?id=20002].

*Hospital recorded diagnoses from the Hospital Episode Statistics (HES) database*

To complement the self-report data assessing HTN and depression above with a clinical evaluation, we used the secondary ICD-10 diagnoses codes a participant has had recorded across all their hospital inpatient records [https://biobank.ndph.ox.ac.uk/showcase/field.cgi?id=41204]. For HES-diagnosed

HTN, we included all occurrences of code I10 ‘Essential (primary) hypertension’. For HES-diagnosed depression, we included all occurrences within codes F32 ‘Depressive episode’ and F33 ‘Recurrent depressive disorder’ without psychotic symptoms. The occurrences were summed per participant and coded as 0 or 1 for the absence or presence of HTN or depression, respectively.

*Supplementary Table 1 – Imaging derived phenotypes used in this study*

| Data field ID | Imaging-Derived Phenotype (IDP) |
|---|---|
| 25054 | Median z-statistic (in group-defined amygdala activation mask) for faces-shapes contrast |
| 25050 | Median z-statistic (in group-defined mask) for faces-shapes contrast |

##### *Multiple Imputation of missing data*

To account for bias due to missing data, we imputed data and conducted sensitivity analyses on the entire sample. Analyses reported in the main manuscript were based on the complete-case data with listwise exclusion of missing values, and the cross-sectional models at baseline assessment were repeated in imputed data for comparison. Data were imputed for individuals without values for predictor, outcome and/or covariates using multiple imputation. Tabulation of missing data showed percentages of missing data below 10% for predictor and outcome variables (Table 1 in main manuscript), except for well-being variables which were introduced at a later timepoint during the baseline assessment (see notes in related data field, e.g. https://biobank.ctsu.ox.ac.uk/crystal/field.cgi?id=4526). Most missing data occurred in covariates for diagnoses of angina, heart attack and lifetime depression (N = 127,467; 24%). To impute datasets, we created 20 imputations using multiple imputation by chained equations with the R package MICE (Van Buuren, 2018; White et al., 2011). Linear regression was performed on each imputed dataset and estimates of the results were then averaged across the imputed datasets in accordance with Rubin’s rules (Rubin, 2004). The results from imputed datasets for the cross-sectional association between mental health and SBP/HTN/number of antihypertensives indicated no sign of bias due to missing data and replicated our results from complete-case analyses (Supplementary Table 2).

*Supplementary Table 2 – Comparison of cross-sectional models derived from analyses using datasets with listwise exclusion of missing data (complete cases) and multiple imputation (imputed). All models included age, sex, BMI, resting heart rate, diabetes diagnosis, lifetime depression diagnosis, angina diagnosis, and myocardial infarction diagnosis as covariates.*

| | Outcome | |
|---|---|---|
| | **Depressive symptoms** | **Well-being** |

| | Predictor | *Estimate* | *SE* | *p* | *Estimate* | *SE* | *p* |
|---|---|---|---|---|---|---|---|
| **Complete cases** | SBP | -0.002 | 0.000 | <0.001 | 0.002 | 0.000 | <0.001 |
| | HTN | 0.049 | 0.003 | <0.001 | -0.070 | 0.004 | <0.001 |
| | No. anti-HTN | -0.005 | 0.001 | 0.001 | 0.007 | 0.002 | 0.005 |
| **Imputed** | SBP | -0.002 | 0.000 | <0.001 | 0.002 | 0.000 | <0.001 |
| | HTN | 0.083 | 0.002 | <0.001 | -0.094 | 0.004 | <0.001 |
| | No. anti-HTN | -0.002 | 0.001 | 0.043 | 0.006 | 0.002 | 0.015 |

#### *Diagnoses and details on medication intake for sensitivity analyses*

*Supplementary Table 3 – List of severe disease diagnoses which were considered in sensitivity analyses. Coding refers to the coding as specified in the source data field f.20002.0*

| Coding | Disease diagnosis |
|---|---|
| 1066 | heart/cardiac problem |
| 1073 | gestational hypertension/pre-eclampsia |
| 1074 | angina |
| 1076 | heart failure/pulmonary odema |
| 1077 | heart arrhythmia |
| 1078 | heart valve problem/heart murmur |
| 1079 | cardiomyopathy |
| 1080 | pericardial problem |
| 1081 | stroke |
| 1082 | transient ischaemic attack (tia) |
| 1083 | subdural haemorrhage/haematoma |
| 1086 | subarachnoid haemorrhage |
| 1093 | pulmonary embolism +/- dvt |
| 1123 | sleep apnoea |
| 1158 | liver failure/cirrhosis |
| 1192 | renal/kidney failure |
| 1193 | renal failure requiring dialysis |
| 1194 | renal failure not requiring dialysis |
| 1196 | urinary tract infection/kidney infection |
| 1200 | ureteric obstruction/hydronephrosis |
| 1224 | thyroid problem (not cancer) |
| 1225 | hyperthyroidism/thyrotoxicosis |
| 1229 | parathyroid gland problem (not cancer) |
| 1230 | parathyroid hyperplasia/adenoma |
| 1232 | disorder of adrenal gland |
| 1233 | adrenal tumour |
| 1235 | hyperaldosteronism/conn's syndrome |
| 1236 | phaeochromocytoma |
| 1237 | disorder or pituitary gland |
| 1238 | pituitary adenoma/tumour |
| 1239 | cushings syndrome |
| 1243 | psychological/psychiatric problem |
| 1244 | infection of nervous system |
| 1245 | brain abscess/intracranial abscess |
| 1246 | encephalitis |
| 1247 | meningitis |
| 1258 | chronic/degenerative neurological problem |
| 1259 | motor neurone disease |
| 1260 | myasthenia gravis |
| 1261 | multiple sclerosis |
| 1262 | parkinsons disease |
| 1263 | dementia/alzheimers/cognitive impairment |
| 1287 | anxiety/panic attacks |
| 1288 | nervous breakdown |
| 1289 | schizophrenia |
| 1290 | deliberate self-harm/suicide attempt |
| 1291 | mania/bipolar disorder/manic depression |
| 1350 | polycystic ovaries/polycystic ovarian syndrome |
| 1371 | sarcoidosis |
| 1372 | vasculitis |
| 1373 | connective tissue disorder |
| 1376 | giant cell/temporal arteritis |
| 1377 | polymyalgia rheumatica |
| 1378 | wegners granulmatosis |
| 1379 | microscopic polyarteritis |
| 1380 | polyartertis nodosa |
| 1381 | systemic lupus erythematosis/sle |
| 1382 | sjogren's syndrome/sicca syndrome |
| 1383 | dermatopolymyositis |
| 1384 | scleroderma/systemic sclerosis |
| 1397 | other demyelinating disease (not multiple sclerosis) |

| | |
|---|---|
| 1405 | other renal/kidney problem |
| 1408 | alcohol dependency |
| 1409 | opioid dependency |
| 1410 | other substance abuse/dependency |
| 1425 | cerebral aneurysm |
| 1426 | myocarditis |
| 1427 | polycystic kidney |
| 1429 | acromegaly |
| 1430 | hypopituitarism |
| 1431 | hyperprolactinaemia |
| 1432 | carcinoid syndrome/tumour |
| 1434 | other neurological problem |
| 1437 | myasthenia gravis |
| 1438 | polycythaemia vera |
| 1445 | clotting disorder/excessive bleeding |
| 1469 | post-traumatic stress disorder |
| 1470 | anorexia/bulimia/other eating disorder |
| 1471 | atrial fibrillation |
| 1480 | dermatomyositis |
| 1481 | polymyositis |
| 1483 | atrial flutter |
| 1484 | wolff parkinson white / wpw syndrome |
| 1485 | irregular heart beat |
| 1486 | sick sinus syndrome |
| 1487 | svt / supraventricular tachycardia |
| 1488 | mitral valve prolapse |

| | |
|---|---|
| 1489 | mitral stenosis |
| 1490 | aortic stenosis |
| 1491 | brain haemorrhage |
| 1506 | primary biliary cirrhosis |
| 1508 | jaundice (unknown cause) |
| 1519 | kidney nephropathy |
| 1520 | iga nephropathy |
| 1546 | essential thrombocytosis |
| 1561 | raynaud's phenomenon/disease |
| 1583 | ischaemic stroke |
| 1584 | mitral valve disease |
| 1585 | mitral regurgitation / incompetence |
| 1586 | aortic valve disease |
| 1587 | aortic regurgitation / incompetence |
| 1588 | hypertrophic cardiomyopathy (hcm / hocm) |
| 1589 | pericarditis |
| 1590 | pericardial effusion |
| 1604 | alcoholic liver disease / alcoholic cirrhosis |
| 1607 | diabetic nephropathy |
| 1608 | nephritis |
| 1609 | glomerulnephritis |
| 1615 | obsessive compulsive disorder (ocd) |
| 1616 | insomnia |
| 1659 | meningioma / benign meningeal tumour |

#### Medication intake

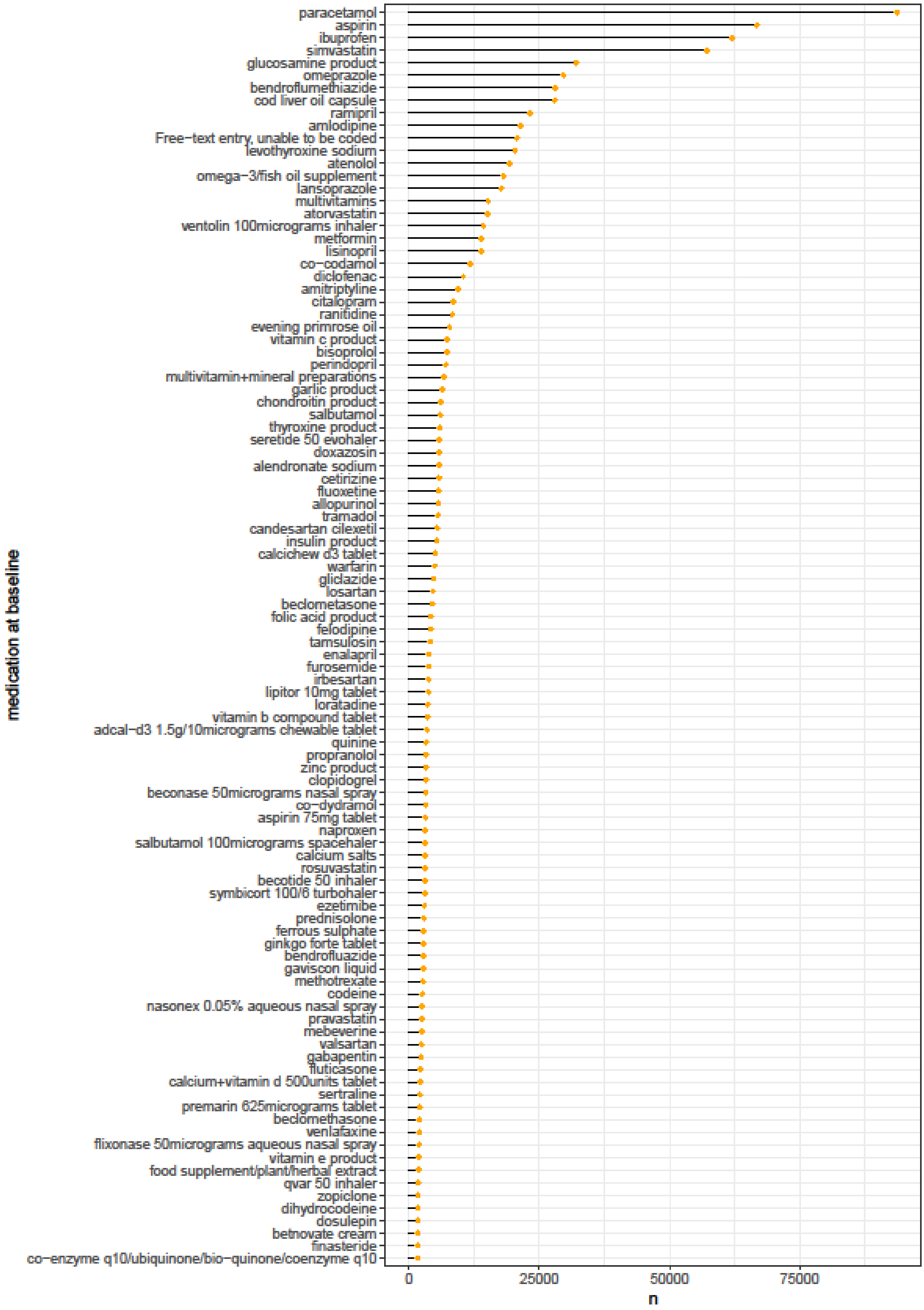


*Supplementary Figure 1 - Medication intake frequency of UK Biobank participants at baseline (100 most frequently reported drugs depicted).*

#### Antihypertensive drug classes

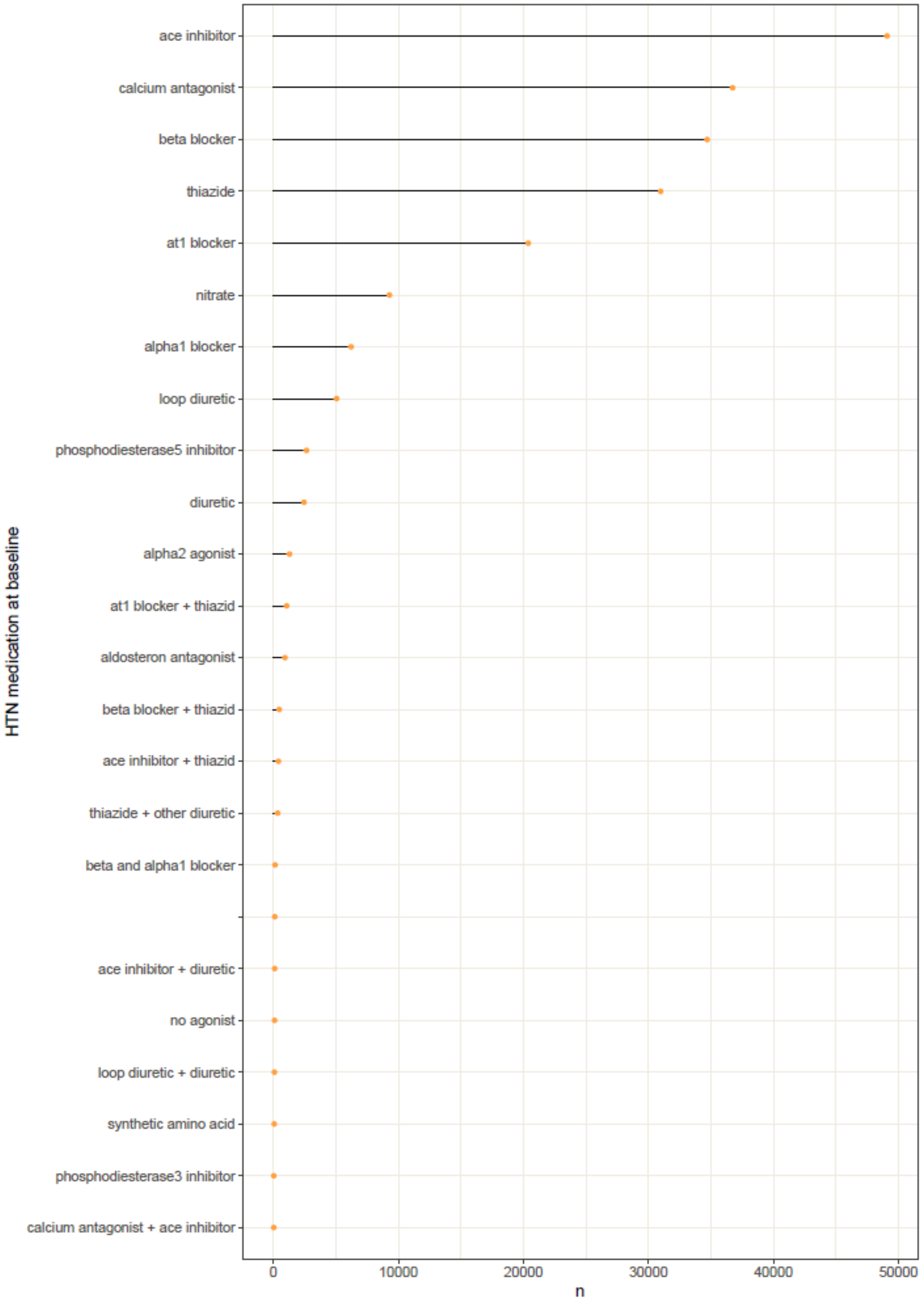


*Supplementary Figure 2 – Frequencies of antihypertensive drug classes at baseline.*

#### Antidepressant drug classes

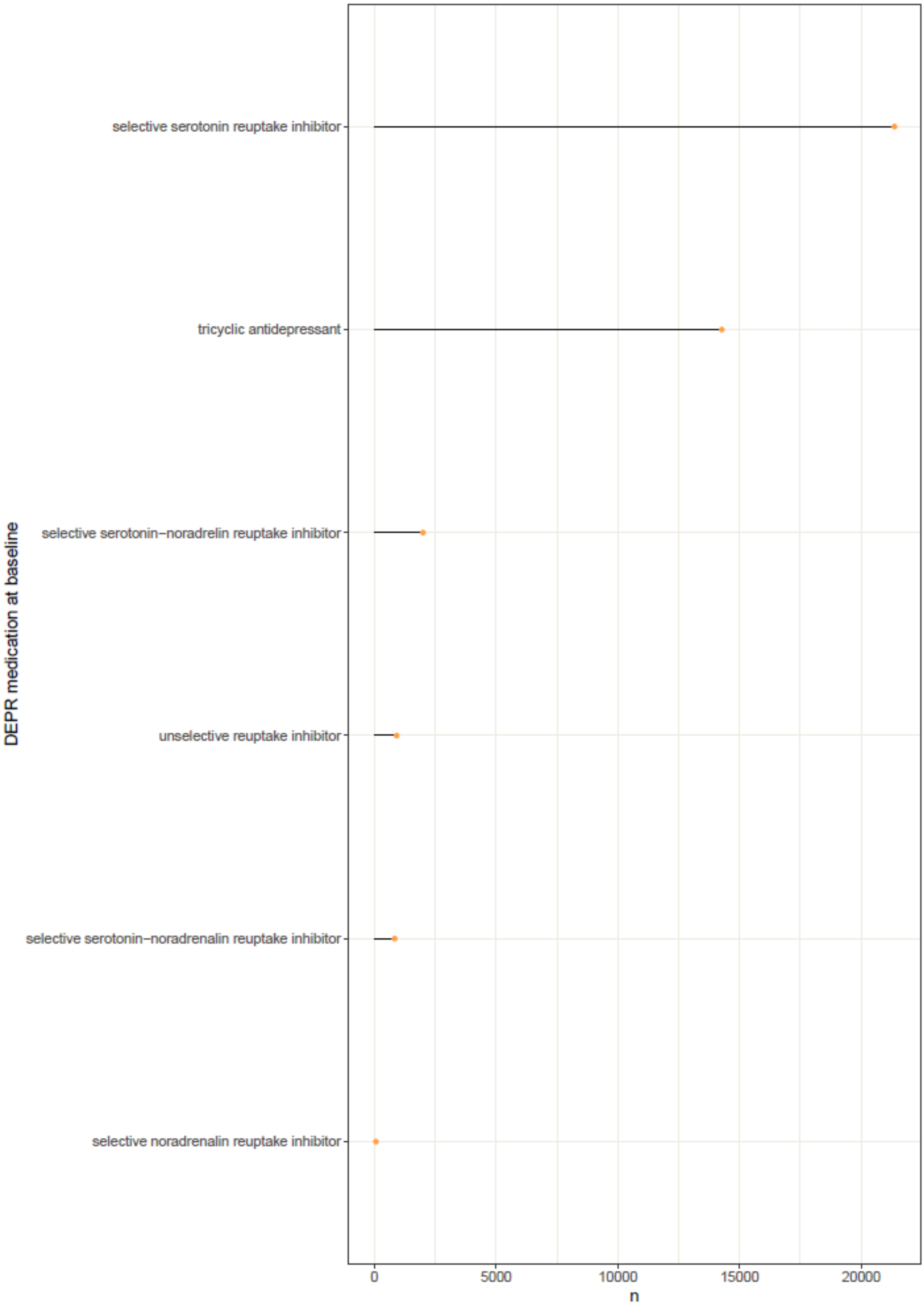


*Supplementary Figure 3 – Frequencies of antidepressant drug classes at baseline.*

#### Supplementary Results

##### *PHQ-9 results*

Longitudinally, we found that baseline HTN was associated with more depressive symptoms assessed with the PHQ-9 approximately 6 years after the initial assessment (β = 0.033; 95% CI [0.025, 0.041]; p<0.001). Similar to the cross-sectional results, higher SBP was related to fewer depressive symptoms at the 6-year-follow-up assessment (PHQ-9: β = -0.049; 95% CI [-0.056, -0.042]; p<0.001). Number of antihypertensive medications at baseline was not significantly associated with the PHQ-9 depression score. The results from the PHQ-9 highly resemble the results using the depression score reported in the main manuscript (Supplementary Figure 4).

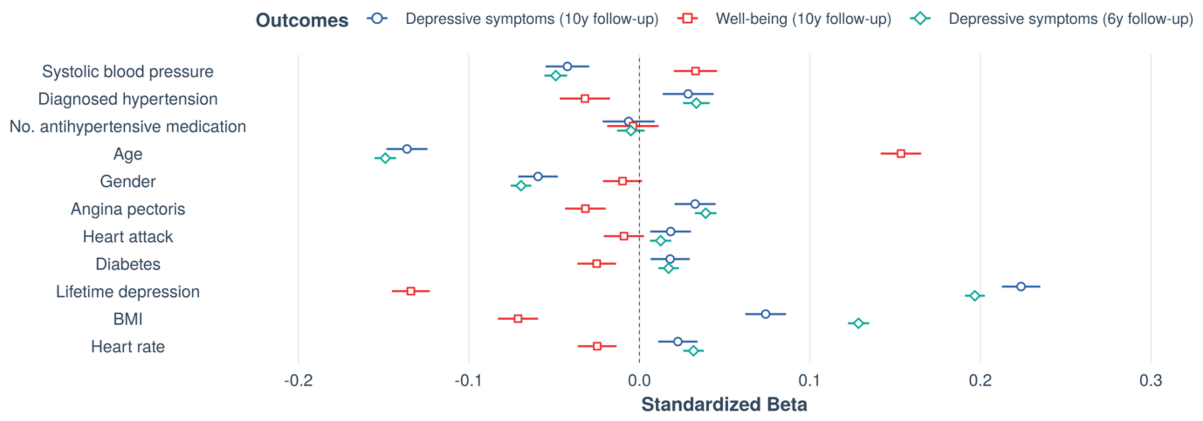


*Supplementary Figure 4 – Longitudinal associations with mental health outcomes including PHQ-9 (i.e., "Depressive symptoms 6y follow-up") at follow-up assessment. Forest plot shows standardized beta estimates and 95% confidence intervals for predictors of interest (systolic blood pressure, diagnosed hypertension (HTN), and number of antihypertensives) as well as covariates at baseline.*

##### *Modelling of additional relevant variables*

Further influencing factors have been previously shown to be important effect modifiers for associations with mental health and/or hypertension, such as insomnia (Kayano et al., 2015), socioeconomic status and education (Kivimäki et al., 2020; Leng et al., 2015), as well as race and ethnic background (Kramer et al., 2004). To assess the robustness of our cross-sectional results to these effects, we analysed an extended version of our cross-sectional model adjusting additionally for insomnia [https://biobank.ndph.ox.ac.uk/ukb/field.cgi?id=1200], household income [https://biobank.ndph.ox.ac.uk/ukb/field.cgi?id=738], educational attainment [https://biobank.ndph.ox.ac.uk/ukb/field.cgi?id=6138], and racial/ethnic background [https://biobank.ndph.ox.ac.uk/ukb/field.cgi?id=21000]. Racial/ethnic background was recoded as a dichotomous variable (White / People of Colour [including people who identified as Asian or Asian British, Black or Black British, Chinese, Mixed, or Other]) as the vast majority of participants selfidentified as white. We also included the UK Biobank assessment centre to model that data acquired at the same assessment centre might be correlated. Among these additionally included covariates (Supplementary Figure 5), insomnia had the largest effect (i.e., standardized beta) on both depressive symptoms (β = 0.224; 95% CI [0.221, 0.228]; $p<0.001$) and well-being (β = -0.172; 95% CI [-0.177, -0.166]; $p<0.001$). While all other additionally included covariates also had some effect on mental health, the effects of our main variables of interest (SBP, HTN, antiHTN) on both mental health outcomes remained virtually unchanged when controlling for all these confounders.

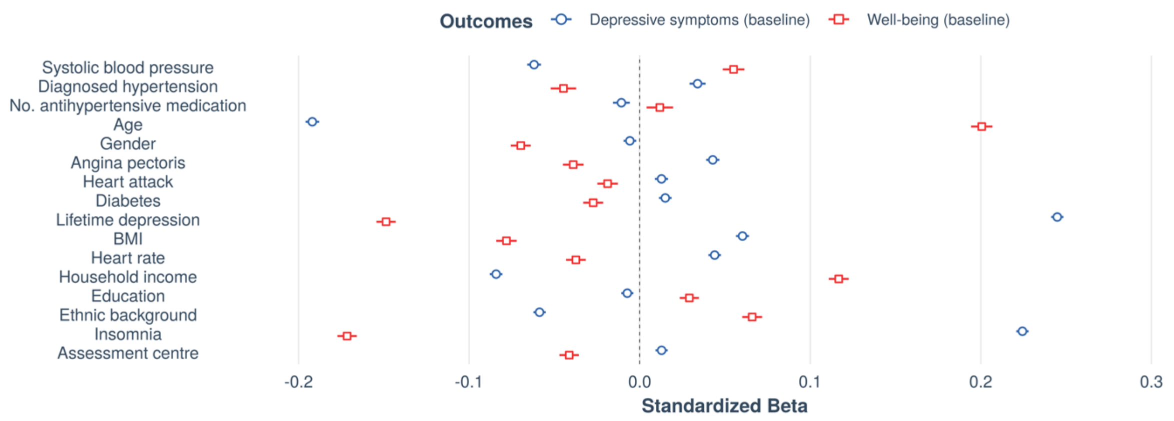


*Supplementary Figure 5 – Cross-sectional associations with mental health outcomes including additional covariates (i.e., insomnia, assessment centre, household income, education and racial/ethnic background) at baseline assessment. Forest plot shows standardized beta estimates and 95% confidence intervals for predictors of interest (systolic blood pressure, diagnosed hypertension (HTN), and number of antihypertensives) as well as covariates at baseline.*

##### *Analyses with hospital records (HES)*

To complement our results using self-report data with clinical assessments, we repeated our analysis using hospital diagnoses for HTN and depression from the HES database. Among the baseline sample (N = 502,494), a total of 340,896 individuals had HES data available. HTN was diagnosed in 112,554 (33%) of HES cases and depression in 18,274 (5%) of HES cases, which shows that HES records may underestimate the prevalence of these conditions in the general population (Davis et al., 2020; Fry et al., 2017). Agreement between self-reported conditions and HES diagnoses was assessed using Cohen's kappa. In line with previous research (Okura et al., 2004), we observed moderate agreement for HTN (kappa = 0.583) and fair agreement for depression (kappa = 0.324).

Next, we used HES-diagnosed HTN to repeat our analysis testing the cross-sectional relationship between depressive symptoms/well-being and SBP, HTN and antihypertensive medication. Including HES-diagnosed HTN instead of self-reported HTN replicated our main results (Supplementary Figure

6). We again observed that SBP was negatively related to depressive symptoms (β = -0.064; 95% CI [-0.068, -0.060]; p<0.001), whereas HTN was related to more depressive symptoms (β = 0.069; 95% CI [0.065, 0.073]; p<0.001) and there was a negative relationship between the number of antihypertensive medications taken and depressive symptoms (β = -0.011; 95% CI [-0.015, -0.006]; p < 0.001). The results yielded larger effect size estimates (i.e., standardized betas) for all these predictors as compared to the analysis with self-reported HTN (Supplementary Figure 6). Inversely, SBP was positively associated with well-being (β = 0.054; 95% CI [0.048, 0.059]; p<0.001), yet the standardized beta coefficient was slightly lower than in the analysis using self-reported HTN. HES HTN related to poorer well-being (β = -0.068; 95% CI [-0.074, -0.061]; p<0.001) with a larger standardized beta coefficient than in the analysis using self-reported HTN. Finally, the relation between antihypertensives and well-being was also not significant in the model including HES HTN (β = 0.004; 95% CI [-0.002, 0.011]; p = 0.193). Overall, the models including HES HTN explained slightly more variance, as reflected in greater adjusted $R^2$-values compared to the models using self-report HTN (depressive symptoms: adj. $R^2$ = 0.131; well-being: adj. $R^2$ = 0.089).

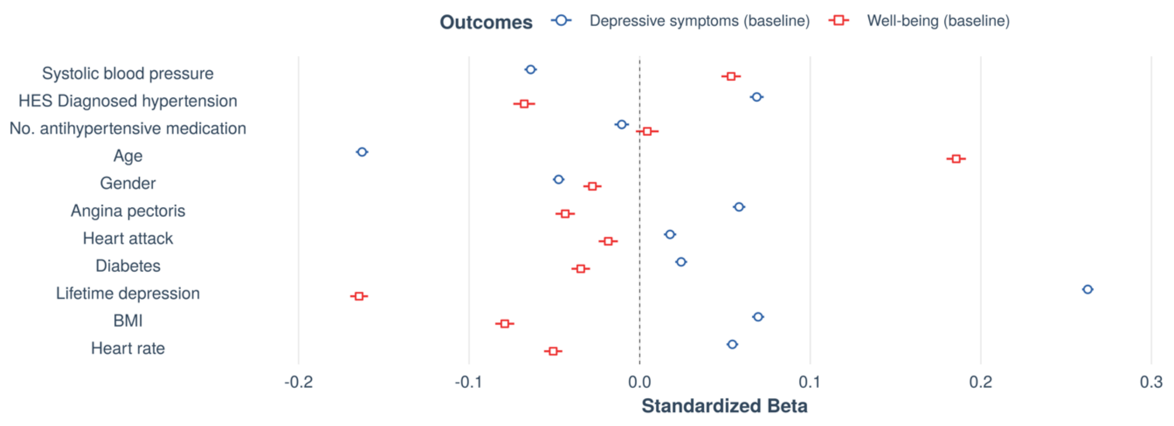


*Supplementary Figure 6 – Cross-sectional associations with mental health outcomes including hospital-diagnosed HTN at baseline assessment. Forest plot shows standardized beta estimates and 95% confidence intervals for predictors of interest (systolic blood pressure, HES diagnosed hypertension, and number of antihypertensives) as well as covariates at baseline.*

Finally, we included an additional analysis testing the relationship of SBP, self-reported HTN and antihypertensive medication with occurrence of HES-diagnosed depression (N=340,900, Supplementary Figure 7). This analysis yielded converging effects with our findings based on self-reports, showing that higher SBP was associated with a lower occurrence of HES-recorded depression (β = -0.040; 95% CI [-0.043, -0.036]; p<0.001), whereas a HTN diagnosis showed a small effect in the opposite direction (β = 0.009; 95% CI [0.005, 0.014]; p<0.001). There was also a small positive association between the number of antihypertensives taken and the occurrence of depression (β = 0.008; 95% CI [0.004, 0.013]; p=0.001). The overall model fit was low (adj. $R^2$ = 0.010).

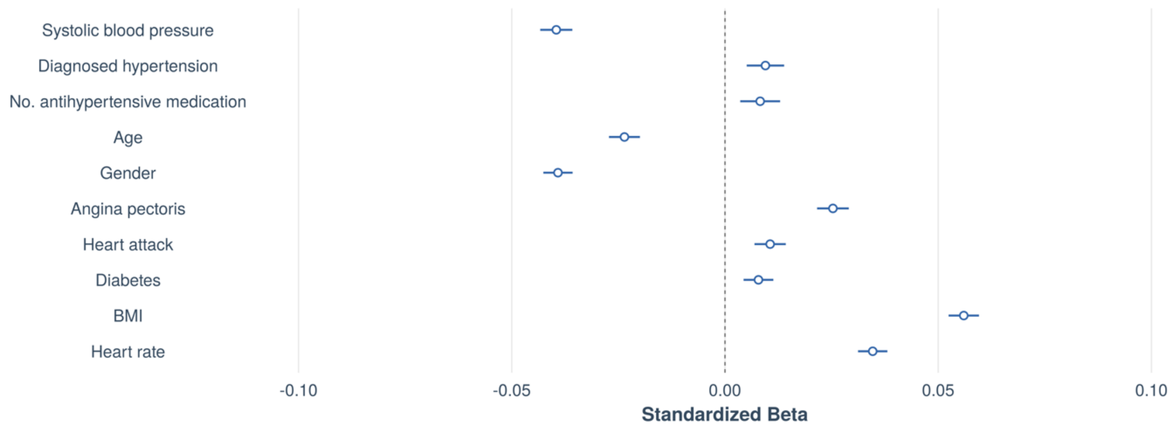


*Supplementary Figure 7 – Cross-sectional associations with hospital-diagnosed depression as outcome. Forest plot shows standardized beta estimates and 95% confidence intervals for predictors of interest (systolic blood pressure, self-reported hypertension diagnosis, and number of antihypertensives) as well as covariates at baseline.*

##### *Assessment of survival bias*

To assess potential survival bias in our sample, we used UK Biobank's death records and observed that 20,442 participants died between the two timepoints of our study, i.e., between the baseline visit and the imaging follow-up (~ 10 years later). The non-surviving sample was older, had higher blood pressure and reported more diagnoses than the total sample (Supplementary Table 4). To assess if there was any survival bias on our results, we investigated our covariate-adjusted cross-sectional model in the non-surviving sample (N = 13,277 for depressive symptoms as outcome; N = 4,865 for well-being as outcome). We again observed that SBP was negatively related to depressive symptoms (β = -0.073; 95% CI [-0.090, -0.056]; $p<0.001$), whereas HTN was related to more depressive symptoms (β = 0.047; 95% CI [0.028, 0.067]; $p<0.001$). There was no significant relationship between the number of antihypertensive medications taken and depressive symptoms (β = -0.004; 95% CI [-0.026, 0.019]; p = 0.749). Inversely, SBP was positively associated with well-being (β = 0.063; 95% CI [0.035, 0.092]; $p<0.001$). The relationship of well-being with HTN (β = -0.016; 95% CI [-0.049, 0.017]; p=0.350) was not significant, while well-being was negatively associated with number of antihypertensives (β = -0.040; 95% CI [-0.078, -0.003]; p = 0.036). The results yielded larger effect size estimates (i.e., standardized betas) for SBP as compared to the analysis in the surviving sample, which could indicate that the effect of SBP on mental health might be particularly relevant for populations at risk with very high SBP levels. Overall, the models in the non-surviving sample explained slightly more variance, as reflected in greater adjusted $R^2$-values compared to the models in the surviving sample (depressive symptoms: adj. $R^2 = 0.132$; well-being: adj. $R^2 = 0.098$).

*Supplementary Table 4 – Sample characteristics at baseline assessment for total sample and non-surviving sub-sample.*

| | Died after baseline (N=20,442) | Overall (N=502,494) |
|---|---|---|
| **Gender** | | |
| Female | 8116 (39.7%) | 273378 (54.4%) |
| Male | 12326 (60.3%) | 229115 (45.6%) |
| Missing | 0 (0%) | 1 (0.0%) |
| **Age (years)** | | |
| Mean (SD) | 61.4 (6.55) | 56.5 (8.10) |
| Median [Min, Max] | 63.0 [40.0, 70.0] | 58.0 [37.0, 73.0] |
| Missing | 0 (0%) | 1 (0.0%) |
| **Townsend deprivation index** | | |
| Mean (SD) | -0.658 (3.42) | -1.29 (3.10) |
| Median [Min, Max] | -1.55 [-6.26, 10.9] | -2.14 [-6.26, 11.0] |
| Missing | 20 (0.1%) | 624 (0.1%) |
| **Systolic blood pressure (mmHg)** | | |
| Mean (SD) | 142 (19.9) | 138 (18.6) |
| Median [Min, Max] | 141 [76.5, 254] | 136 [65.0, 254] |
| Missing | 2307 (11.3%) | 45540 (9.1%) |
| **Diastolic blood pressure (mmHg)** | | |
| Mean (SD) | 82.1 (10.8) | 82.2 (10.1) |
| Median [Min, Max] | 82.0 [36.5, 133] | 82.0 [36.5, 148] |
| Missing | 2305 (11.3%) | 45528 (9.1%) |
| **Heart rate (beats/min)** | | |
| Mean (SD) | 72.1 (12.8) | 69.3 (11.2) |
| Median [Min, Max] | 71.0 [33.5, 148] | 68.5 [30.5, 173] |
| Missing | 2305 (11.3%) | 45528 (9.1%) |
| **BMI (kg/m2)** | | |
| Mean (SD) | 28.2 (5.44) | 27.4 (4.80) |
| Median [Min, Max] | 27.4 [12.8, 74.7] | 26.7 [12.1, 74.7] |
| Missing | 328 (1.6%) | 3105 (0.6%) |
| **Diabetes** | | |
| Prefer not to answer | 21 (0.1%) | 404 (0.1%) |
| Do not know | 81 (0.4%) | 1280 (0.3%) |
| No | 17592 (86.1%) | 473479 (94.2%) |
| Yes | 2700 (13.2%) | 26399 (5.3%) |
| Missing | 48 (0.2%) | 932 (0.2%) |
| **Angina** | | |
| No diagnosed angina or unknown | 15513 (75.9%) | 358910 (71.4%) |
| Diagnosed angina | 1883 (9.2%) | 16117 (3.2%) |
| Missing | 3046 (14.9%) | 127467 (25.4%) |
| **Heart attack** | | |
| No diagnosed heart attack or unknown | 15762 (77.1%) | 363524 (72.3%) |
| Diagnosed heart attack | 1634 (8.0%) | 11503 (2.3%) |
| Missing | 3046 (14.9%) | 127467 (25.4%) |
| **Lifetime depression** | | |
| No diagnosed depression or unknown | 16028 (78.4%) | 346919 (69.0%) |
| Diagnosed depression | 1368 (6.7%) | 28108 (5.6%) |

| | | |
|---|---|---|
| Missing | 3046 (14.9%) | 127467 (25.4%) |
| **No. antihypertensive medication** | | |
| Mean (SD) | 0.852 (1.25) | 0.403 (0.864) |
| Median [Min, Max] | 0 [0, 9.00] | 0 [0, 9.00] |
| **No. antidepressant medication** | | |
| Mean (SD) | 0.120 (0.346) | 0.0784 (0.281) |
| Median [Min, Max] | 0 [0, 3.00] | 0 [0, 5.00] |
| **Current depressive symptoms** | | |
| Mean (SD) | 1.48 (0.599) | 1.40 (0.528) |
| Median [Min, Max] | 1.25 [1.00, 4.00] | 1.25 [1.00, 4.00] |
| Missing | 2662 (13.0%) | 53563 (10.7%) |
| **Well-being** | | |
| Mean (SD) | 4.37 (0.620) | 4.46 (0.579) |
| Median [Min, Max] | 4.40 [1.00, 6.00] | 4.50 [1.00, 6.00] |
| Missing | 14704 (71.9%) | 330042 (65.7%) |
| **Diagnosed hypertension** | | |
| No diagnosed HTN or unknown | 12070 (59.0%) | 365819 (72.8%) |
| Diagnosed HTN | 8324 (40.7%) | 135745 (27.0%) |
| Missing | 48 (0.2%) | 930 (0.2%) |

##### *Moderation analysis of BP-mental health relationship with SBP as additional criterion to define hypertension status*

In the moderation analysis (Figure 4 in main manuscript), we observed that among people who were regarded as "normotensive" based on the two criteria "lack of a previous HTN diagnosis" and "no intake of antihypertensive medication", there were some individuals with baseline blood pressure levels of systolic 140 mmHg or higher. We therefore refined our criteria to thoroughly detect all hypertensive participants at both initial assessment (for conservative exclusion) and at follow-up (to clearly define who will develop hypertension). We thus repeated the moderation analysis while considering SBP >140 mmHg as an additional criterion (next to HTN diagnosis and antihypertensive medication) for the definition of hypertension. Thus, we excluded all participants who were hypertensive at the initial assessment (defined as SBP>140 mmHg, HTN diagnosis or intake of antihypertensives), leaving us with a sample of N=17,879 participants with data at baseline and follow-up. In this sample of non-hypertensive participants, mean baseline SBP was 124 mmHg (SD = 9.92). Among these, those who stayed normotensive until follow-up had a mean SBP of 122 mmHg (SD = 10.0) and those who developed HTN had a mean SBP of 129 mmHg (SD = 7.76). Across all non-hypertensive participants, SBP increased significantly from initial assessment to follow-up (mean increase = 6.885 mmHg; $t$ = 67.432; degrees of freedom = 17878; 95% CI [6.686, 7.086]; $p$<0.001). Within this sample, we replicated the results obtained in the larger group: In unadjusted models, there were no significant group differences in mental health at initial assessment between people who developed HTN and those who stayed normotensive (HTN: mean depressive symptoms = 1.347; no HTN: mean depressive symptoms = 1.358; $t$ = 1.346; degrees of freedom = 9608; 95% CI [-0.005, 0.026]; $p$=0.178; HTN: mean well-being = 4.525; no HTN: mean well-being = 4.523; $t$ = -0.109; degrees of freedom = 2888.2; 95% CI [-0.033, 0.029]; $p$=0.913). However, in the fully adjusted regression model (e.g., adjusting for baseline blood pressure), we again observed a main effect of later HTN on baseline mental health (depressive symptoms: $\beta$ = 0.038; 95% CI [0.021, 0.056]; $p$<0.001; well-being: $\beta$ = -0.029; 95% CI [-0.058, 0.001]; $p$=0.056), suggesting that when adjusting for SBP levels, people who later developed HTN reported more depressive symptoms already at initial assessment compared to people without HTN. We also replicated that the negative association between depressive symptoms and SBP at initial assessment was moderated by HTN at follow-up ($\beta$ = -0.021; 95% CI [-0.039, -0.004]; p=0.019). The moderation was however not significant for depressive symptoms at follow-up. Neither did we observe any significant moderation effects of developing HTN for well-being at any of the assessments. In sum, these results corroborate our findings that people who developed HTN throughout the course of the study might require higher SBP levels to sustain the same mental health outcomes as normotensives and that the negative relationship between mental health and blood pressure was accentuated in people developing HTN several years before HTN manifested.

##### *Inclusion/exclusion of previous depression and other BP-altering diseases*

We also tested if any effects of SBP, HTN and antihypertensive intake on current depressive symptoms and well-being depended on the presence or absence of previous diagnosis of depression or any other severe disease that might affect BP (list of diseases in Supplementary Table 3).

Conducting the analyses separately in groups of participants who either have had no or any previous diagnosis of depression yielded similar cross-sectional results as in the total sample (all p<0.001, Supplementary Table 5). As described in Supplementary Table 5, in people both with or without depression diagnosis, SBP was negatively associated with depressive symptoms and positively with well-being. Inversely, in both sub-groups HTN was positively related with depressive symptoms and negatively with well-being. Higher intake of antihypertensives was related to fewer depressive symptoms only in people without previous depression.

The results were also robust in sub-samples of people without or with diseases which often co-occur with changes in blood pressure, such as cardiovascular or renal diseases (Supplementary Table 5). Only number of antihypertensives yielded diverging results between these sub-groups: While people without disease diagnoses, showed a negative association between number of antihypertensives and depressive symptoms and a positive one with well-being scores, higher intake of antihypertensives was related to more depressive symptoms and lower well-being in people with any diseases (Supplementary Table 5).

*Supplementary Table 5 – Sensitivity analyses with inclusion and exclusion of subsamples with disease diagnoses at initial assessment.*

| | Sub-group | *N* | SBP (β) | HTN (β) | No. anti-HTN (β) | Adj. $R^2$ |
|---|---|---|---|---|---|---|
| **Depressive symptoms** | Depression diagnosis | 22,438 | -0.052 | 0.045 | n.s. | 0.066 |
| | No depression diagnosis | 281,333 | -0.069 | 0.046 | -0.008 | 0.052 |
| | Any diagnosis | 59,126 | -0.055 | 0.037 | 0.028 | 0.149 |
| | No diagnosis | 244,645 | -0.063 | 0.050 | -0.022 | 0.119 |
| **Well-being** | Depression diagnosis | 9,140 | 0.059 | -0.082 | n.s. | 0.103 |
| | No depression diagnosis | 120,736 | 0.058 | -0.055 | n.s. | 0.053 |
| | Any diagnosis | 24,946 | 0.046 | -0.045 | -0.024 | 0.116 |
| | No diagnosis | 104,930 | 0.058 | -0.064 | 0.021 | 0.078 |

#### *Medication effects*

A major characteristic of the UK Biobank sample is the large number of people taking any kind of medication (*N* = 363,967). Among the most frequently used medications were pain killers (e.g. paracetamol, *N* = 93,619), antihypertensives (e.g. bendroflumethiazide, *N* = 28,000) and also antidepressants (*N* = 37,699). We therefore explored potential effects of these medications on the relationship between blood pressure and mental health.

##### *Antidepressants*

Sub-group analyses of participants taking or not taking antidepressant medication, showed similar associations between SBP, HTN and mental health as analyses on the total sample (Supplementary Table 6). However, when considering only participants not taking antidepressants, higher intake of antihypertensives was associated with fewer depressive symptoms (β = -0.013; 95% CI [-0.018, -0.008]; $p<0.001$) and greater well-being (β = 0.009; 95% CI [0.002, 0.017]; $p = 0.018$).

##### *Other medications*

We also compared the blood pressure-mental health relationship between participants taking any sort of medications and those who took none (Supplementary Table 6). Again, we found a negative effect of SBP and a positive effect of HTN on depressive symptoms (and the inverse pattern for well-being) in both sub-groups (Supplementary Table 6). Despite the smaller sample sizes, standardized beta estimates were slightly larger for the sub-group of participants who did not take any medications, compared to the total sample (Supplementary Table 6 and see results in main manuscript).

*Supplementary Table 6 – Sensitivity analyses exploring medication effects on blood pressure-mental health associations at initial assessment.*

| | Sub-group | N | SBP (β) | HTN (β) | No. anti-HTN (β) | Adj. $R^2$ |
|---|---|---|---|---|---|---|
| **Depressive symptoms** | Antidepressants | 28,722 | -0.042 | 0.045 | n.s. | 0.110 |
| | No antidepressants | 275,049 | -0.064 | 0.046 | -0.013 | 0.065 |
| | Any medication | 252,073 | -0.057 | 0.027 | - | 0.138 |
| | No medication | 51,698 | -0.080 | 0.068 | - | 0.063 |
| **Well-being** | Antidepressants | 12,046 | 0.040 | -0.068 | n.s. | 0.120 |
| | No antidepressants | 117,830 | 0.056 | -0.057 | 0.009 | 0.059 |
| | Any medication | 105,800 | 0.053 | -0.041 | - | 0.094 |
| | No medication | 24,076 | 0.067 | -0.076 | - | 0.056 |

*Specificity of antidepressant classes*

We conducted follow-up analyses on these effects to explore if specific classes of antidepressants were particularly associated with the mental health measures. With regards to depressive symptoms, we found negative associations with tricyclic antidepressants (estimate = -0.296; 95% CI [-0.558, -0.033]; p=0.027) and negative associations with unselective reuptake inhibitors (estimate = -0.354; 95% CI [-0.623, -0.084]; p=0.010). Associations between well-being and antidepressant classes were not significant (all p>0.05).

*Specificity of antihypertensive classes*

We did not observe any significant effects of antihypertensive classes on depressive symptoms (all p>0.05). Well-being was negatively associated with aldosterone antagonists (estimate = -0.261; 95% CI [-0.502, -0.020]; p=0.034) and positively with the intake of a combination of beta and alpha1 blockers (estimate = 0.347; 95% CI [0.029, 0.664]; p=0.032).

##### *Exploration of unmeasured confounding bias with the E-value*

To also explore the effect of potential unmeasured confounders on the results, we calculated E-values using the "EValue" package in R (VanderWeele & Ding, 2017). E-values indicate the minimum strength of association, on the risk ratio scale, that an unmeasured confounder would need to have with both the exposure and outcome to explain away an exposure–outcome association, above and beyond the measured covariates. For continuous exposure variables, such as SBP in our study, the function uses a dichotomisation in the exposure between hypothetical groups of participants per one-unit increase. For the continuous outcome, such as depressive symptoms and well-being in our study, the function uses an effect-size conversion to approximately convert the mean difference between the exposure groups to the odds ratio that would arise from dichotomising the continuous outcome (VanderWeele & Ding, 2017). As a result, we observed for the association of SBP with depressive symptoms values of E = 1.32 and with well-being of E = 1.30. For the association of HTN with depressive symptoms, we observed E = 1.25 and with well-being E = 1.30. The interpretation of the E-value depends on the context regarding the measured confounders. Given that we have included several plausible confounders in our models, the resulting E-values give further support for the robustness of our results, as an unmeasured confounder would need to be associated with outcome and exposure by a risk ratio of around 1.3-fold each, above and beyond the measured confounders. However, we also note that while E-values have been introduced as robustness analyses for studies aiming to estimate causal effects (VanderWeele & Ding, 2017), we consider them an additional sensitivity measure for robustness. Potential causal effects should more directly be investigated in future studies with experimental designs or observational designs with dedicated causal identification strategies (e.g., directed acyclic graphs).
